# Delayed viral clearance and exacerbated airway hyperinflammation in hypertensive COVID-19 patients

**DOI:** 10.1101/2020.09.22.20199471

**Authors:** Saskia Trump, Soeren Lukassen, Markus S. Anker, Robert Lorenz Chua, Johannes Liebig, Loreen Thürmann, Victor Corman, Marco Binder, Jennifer Loske, Christina Klasa, Teresa Krieger, Bianca P. Hennig, Marey Messingschlager, Fabian Pott, Julia Kazmierski, Sven Twardziok, Jan Philipp Albrecht, Jürgen Eils, Sara Hadzibegovic, Alessia Lena, Bettina Heidecker, Christine Goffinet, Florian Kurth, Martin Witzenrath, Maria Theresa Völker, Sarah Dorothea Müller, Uwe Gerd Liebert, Naveed Ishaque, Lars Kaderali, Leif-Erik Sander, Sven Laudi, Christian Drosten, Roland Eils, Christian Conrad, Ulf Landmesser, Irina Lehmann

## Abstract

In COVID-19, hypertension and cardiovascular diseases have emerged as major risk factors for critical disease progression. Concurrently, the impact of the main anti-hypertensive therapies, angiotensin-converting enzyme inhibitors (ACEi) and angiotensin receptor blockers (ARB), on COVID-19 severity is controversially discussed. By combining clinical data, single-cell sequencing data of airway samples and *in vitro* experiments, we assessed the cellular and pathophysiological changes in COVID-19 driven by cardiovascular disease and its treatment options. Anti-hypertensive ACEi or ARB therapy, was not associated with an altered expression of SARS-CoV-2 entry receptor *ACE2* in nasopharyngeal epithelial cells and thus presumably does not change susceptibility for SARS-CoV-2 infection. However, we observed a more critical progress in COVID-19 patients with hypertension associated with a distinct inflammatory predisposition of immune cells. While ACEi treatment was associated with dampened COVID-19-related hyperinflammation and intrinsic anti-viral responses, under ARB treatment enhanced epithelial-immune cell interactions were observed. Macrophages and neutrophils of COVID-19 patients with hypertension and cardiovascular comorbidities, in particular under ARB treatment, exhibited higher expression of *CCL3, CCL4*, and its receptor *CCR1*, which associated with critical COVID-19 progression. Overall, these results provide a potential explanation for the adverse COVID-19 course in patients with cardiovascular disease, i.e. an augmented immune response in critical cells for the disease course, and might suggest a beneficial effect of clinical ACEi treatment in hypertensive COVID-19 patients.

## Introduction

The ongoing coronavirus disease 2019 (COVID-19) pandemic is caused by the severe acute respiratory syndrome coronavirus 2 (SARS-CoV-2). Among patients hospitalized for COVID-19 males and those of older age emerged as having a higher risk for critical COVID-19^1, 2^. Hypertension, highly prevalent in adults worldwide^3^, has been identified as a major risk factor for COVID-19 severity^4, 5^. Hypertensive COVID-19 patients are more likely to develop severe pneumonia or organ damage than non-hypertensive patients. In addition, these patients exhibit exacerbated inflammatory responses and have a higher risk of dying from COVID-19 than non-hypertensive patients^4, 6^.

SARS-CoV-2 exploits the ACE2 receptor, expressed on epithelial cells in the respiratory system, for cellular attachment and entry^7^. ACE2 is a membrane-bound aminopeptidase and is part of the non-canonical arm of the renin-angiotensin-aldosterone system (RAAS), which regulates blood pressure homeostasis and vascular repair responses. It has been speculated that antihypertensive treatment by angiotensin-converting enzyme inhibitors (ACEi) or angiotensin receptor blockers (ARBs) might modulate ACE2 expression and thereby alter susceptibility for SARS-CoV-2 infection. In the classical RAAS pathway, angiotensin II binds to the angiotensin-II-receptor-subtyp-1 (AT1R), which promotes vasoconstriction and pro-inflammation. ACE2, on the other hand, cleaves angiotensin II into angiotensin 1-7, which mediates vasodilatatory and anti-inflammatory effects^8, 9^.

Data from animal studies demonstrated that ACEi and ARB can up-regulate *ACE2* expression^10^. This raised intense discussions on a potential increase in availability of SARS-CoV-2 receptors in ACEi or ARB treated patients^11, 12^, rendering them potentially more susceptibility to viral infection and spread. To date there is no evidence from observational studies that ACEi-or ARB-treatment could increase the infectivity for SARS-CoV-2^5, 13^.

Hypertension is associated with the activation of inflammatory processes^14, 15^. As a hyperinflammatory phenotype in the respiratory system has been described to enhance severity of COVID-19^16, 17^, we assessed whether a potential pro-inflammatory predisposition of hypertensive patients before SARS-CoV-2 infection may contribute to an exacerbated disease severity.

To this end we evaluated the impact of coexisting cardiovascular illnesses, in particular of hypertension and anti-hypertensive treatment, on COVID-19 pathology and viral clearance based on two German prospective cohorts. By analyzing the single-cell transcriptome landscape of the airways of COVID-19 patients and SARS-CoV-2 negative controls we provide insights into the differential COVID-19 pathology in ACEi/ARB-treated patients compared to those without cardiovascular diseases or different anti-hypertensive treatment.

## Results

### ACEi/ARB treatment was associated with a lower hypertension-related risk for critical COVID-19

We first assessed the impact of hypertension (HT+) and other cardiovascular diseases (CVD+) with anti-hypertensive treatment on COVID-19 severity (Figure 1a). Both medical conditions have been associated with a worse outcome in COVID-19^5, 11-13, 18-20^. Accordingly, we compared the proportion of critical cases to all other severities of COVID-19 in the different patient groups of the Pa-COVID-19 cohort^21^ (see Supplementary Table 1 for clinical characteristics). The proportion of patients with a critical outcome was significantly increased for HT+/CVD± patients (n=90) compared to HT-/CVD-COVID-19 patients (n=54, p-value=0.007). For HT+ patients, the risk for critical COVID-19 was highest without ACEi- or ARB-treatment (see Supplementary Table 2): almost 75% of HT+/CDV-patients and about two thirds of HT+/CVD+ patients showed critical COVID-19. In contrast, ACEi- and ARB-treatment were associated with a decreased proportion of critical COVID-19 in both groups (HT+/CVD- and HT+CVD+), but ACEi treatment showed a more profound difference as compared to ARB treatment. ACEi treated HT+/CVD± patients showed almost the same risk for critical COVID-19 as HT-/CVD-patients (Supplementary Table 2).

**Figure 1.**
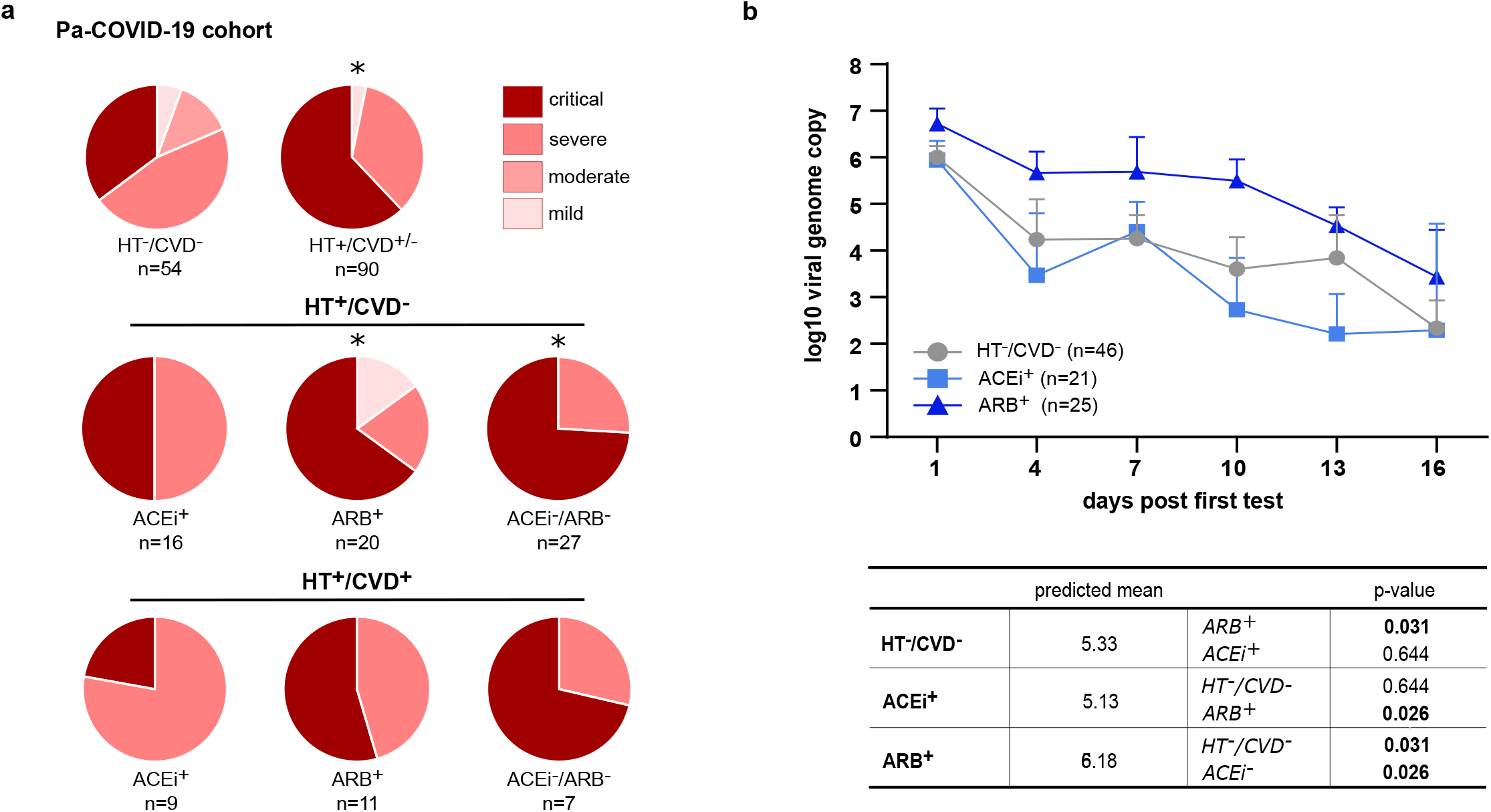
Association of anti-hypertensive treatment with COVID-19 severity and viral clearance. (a) Comparison of COVID-19 severity based on WHO-classification in patients without cardiovascular comorbidities (HT-/CVD-, n=54) and those with arterial hypertension and cardiovascular disease (HT+/CVD±, n=90). CVD patients were separated in those with HT only (HT+/CVD-, n=63) and those with additional cardiovascular diseases (HT+/CVD+, n=27) and are depicted dependent on their treatment with ARB (ARB+), ACEi (ACEi+) or other medications (ACEi-/ARB-). ∗p-value <0.05 based on a Yates-corrected chi-square comparison of critical vs. all other WHO categories. (b) Viral clearance over time shown for SARS-CoV-2 positive patients without a pre-existing cardiovascular disease (CVD-/HT-, n=46) in comparison to ARB+ (n=25) or ACEi+ (n=21) HT+/CVD± COVID-19 patients. Depicted are mean +/- SD of qPCR data binned in 3-day intervals, only the maximal value of each patient in this interval was considered. Adjusted regression analysis (confounder: BMI, gender, smoking, insulin-treatment, days post onset of symptoms, n=92) showed a significantly higher viral load for ARB+/ HT+/CVD±, compared to ACEi+/ HT+/CVD± and HT-/CVD-patients.

To exclude the impact of other risk factors for an adverse COVID-19 clinical course, we performed logistic regression analyses adjusted for known confounding factors such as age, sex, and BMI. This analysis confirmed a higher risk for developing critical COVID-19 for HT+/CVD-over HT-/CVD- (adjOR=2.38, 95%CI:1.09-5.21, p-value=0.028). Even after adjustment for confounders ARB-treatment showed an increased risk for critical COVID-19 (HT+/CVD- /ARB+, adjOR=3.85, 95%CI:1.01-14.68, p-value=0.044), which was lower than for HT+/CVD-patients without ACEi or ARB treatment (adjOR=4.94, 95%CI:1.46-16.79, p-value=0.009). The logistic regression analysis revealed no significant increase for critical COVID-19 by ACEi treatment compared to HT-/CVD-.

Our results showed that patients with hypertensive disease had an increased risk for critical COVID-19. This risk was decreased by ACEi/ARB treatment. ACEi treatment almost entirely abolished the additional risk, whereas ARB treatment only reduced the hypertension-associated risk.

### ARB but not ACEi treatment was associated with delayed SARS-CoV-2 clearance

We investigated the dynamics of SARS-CoV-2 clearance in patients included in the Pa-COVID-19 cohort. During hospitalization, COVID-19 patients were tested longitudinally for SARS-CoV-2 by qPCR of the viral genome. Using an adjusted repeated measurement mixed model, we studied the changes of the viral load over time, comparing ACEi+ (n=21) or ARB+ (n=26) COVID-19 patients with HT-/CVD-COVID-19 patients (n=46). All three groups showed the same initial viral load while ACEi+ treatment did not change viral clearance up to 16 days after the first positive test compared to HT-/CVD-, ARB-treatment was associated with a significantly slower viral clearance over time compared to HT-/CVD- (p-value=0.031) or ACEi+ (p-value= 0.026, Figure 1b), respectively. This finding was supported by the time-dependent slope of viral load between the different patient groups. ARB+ patients tended to have a flatter slope compared to HT-/CVD- (p-value=0.07, Extended Data Figure 1a). The same was observed for HT+/ CVD± patients who showed a tendency of slower viral clearance compared to HT-/CVD-patients (p-value=0.08, Extended Data Figure 1b). Taken together, we showed that viral clearance in HT+/CVD± patients under ACEi treatment was similar to that in COVID-19 patients without a coexisting cardiovascular disease, while viral clearance may have been delayed in patients undergoing hypertensive ARB treatment.

### Cardiovascular disease and SARS-CoV-2 infection affect cell type distribution

To investigate the cellular and molecular impact of cardiovascular co-morbidities and anti-hypertensive treatment on COVID-19 severity, we performed extensive single-cell transcriptome profiling of nasopharyngeal samples from COVID-19 patients with or without hypertension and other cardiovascular diseases (see Supplementary Table 3 for clinical characteristics). To disentangle the effect of HT and CVD on SARS-CoV-2 infection, we studied a mirror cohort of SARS-CoV-2 negative patients with and without HT/CVD under ARB/ACEi treatment (Figure 2a). In total, we assessed the transcriptomes of 114,761 individual cells obtained from nasopharyngeal swabs of 32 COVID-19 patients (n=25 with HT+/CVD±; n=10 ACEi+ and n=15 ARB+) and 16 SARS-CoV-2-negative controls (n=10 with HT+/CVD±; n=6 ACEi+ and n=4 ARB+).

**Figure 2.**
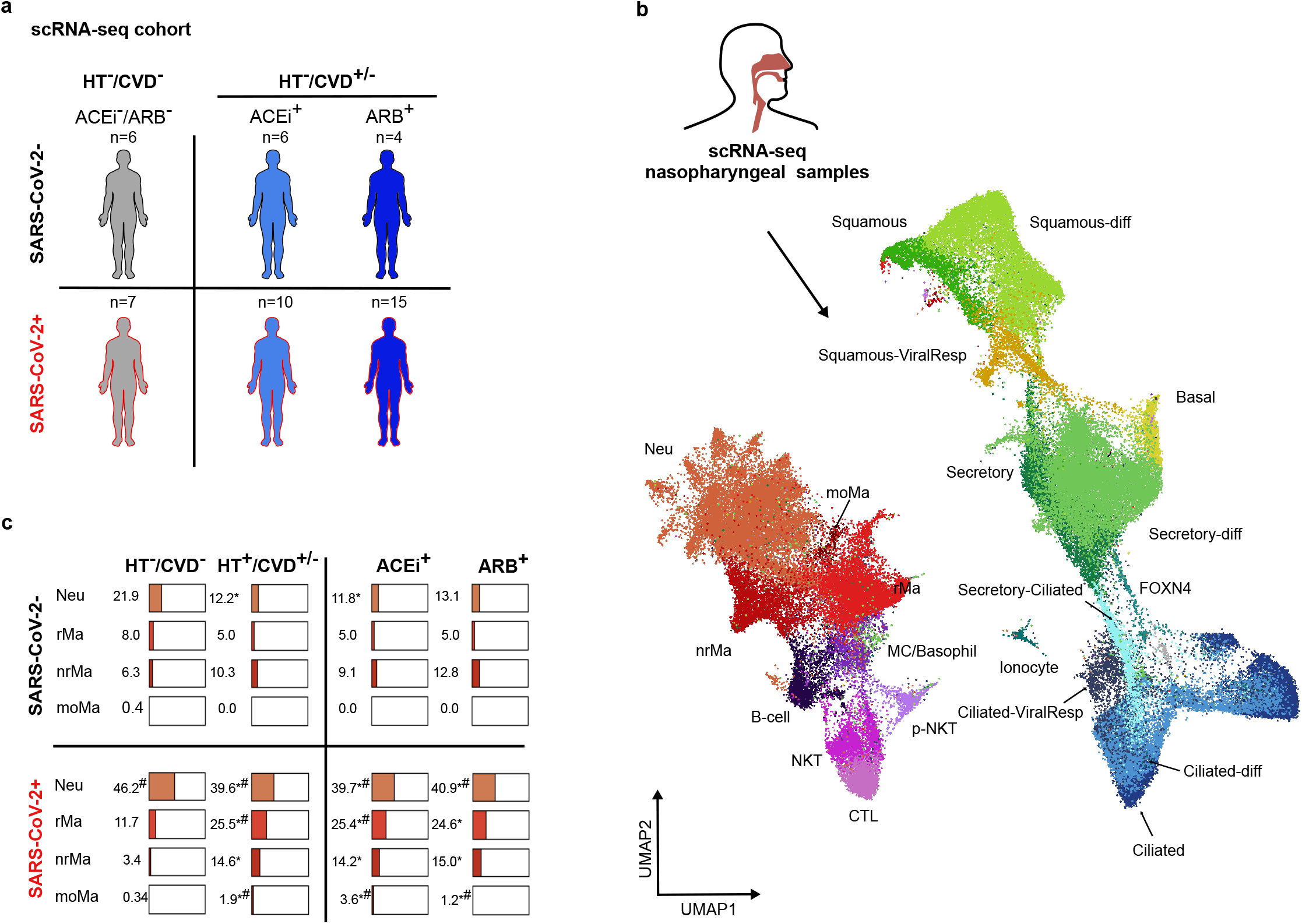
Characteristics of the scRNA-seq cohorts and cell type distribution of nasopharyngeal samples. (a) COVID-19 patients were analysed by scRNA-seq to study the impact of HT/CVD and its treatment by ARB or ACEi in SARS-CoV-2 negative (n=16) and positive patients (n=32). (b) Samples were collected from the nasopharynx of the patients and subjected to scRNA-seq resulting in the given UMAP displaying all identified cell types and states (color-coded). (c) Distribution of selected immune cell types/states in SARS-CoV-2 negative and positive patients separated by HT+/CVD± / HT-/CVD- or ACEi+/ARB+ treatment. Given are percentages related to the total number of immune cells. MC = mast cells; moMa = monocyte derived macrophage; Neu = neutrophil; (n)rMa = (non-)resident macrophage; p-NKT = proliferating natural killer T-cell; CTL = cytotoxic T lymphocyte. ∗significance compared to CVD-/HT-, ^#^ significance compared to SARS-CoV-2 neg.

Only individuals diagnosed with severe to critical COVID-19 or SARS-CoV-2-negative controls were eligible for inclusion in this part of the study (Supplementary Table 3). We identified nine immune and 12 epithelial cell populations (Figure 2b, Extended Data Figure 2 and 3). While the relative abundance of the individual epithelial cell types seemed largely unchanged (Extended Data Figure 3b) the immune cell population upon infection was characterized by a massive increase of neutrophils (Neu, p-value < 2.66e-10) and resident macrophages (rMa, p-value < 5.78e-11, Figure 2c) upon infection. In HT+/CVD± COVID-19 patients, non-resident macrophages (nrMa, p-value <2.10e-14) and monocyte-derived macrophages (moMa, p-value<4.83e-10) were expanded compared to HT-/CVD-patients, independent of their anti-hypertensive treatment (Figure 2c, Extended Data Figure 3b).

### Anti-hypertensive treatment is not associated with altered expression of the SARS-CoV-2 entry receptor *ACE2*

SARS-CoV-2 enters the human cell via the receptor ACE2 and with the help of the protease TMPRSS2. It has been speculated that ARB and ACEi as RAAS-modulating agents might change *ACE2* expression and thereby the infectivity for SARS-CoV-2. Since the expression of *ACE2* is generally low in human airways^22^, we quantified total *ACE2* expression per sample. In line with previous studies^23, 24^, we found an overall increased expression of both *ACE2* (p=0.0025, Extended Data Figure 4a) and *TMPRSS2* (p-value= 0.0002, Extended Data Figure 4b) upon SARS-CoV-2 infection. However, anti-hypertensive treatment did not alter *ACE2* expression, neither in SARS-CoV-2-positive nor -negative patients.

We conclude that entry factor expression did not predispose ACEi or ARB treated patients to SARS-CoV-2 infection. This finding is in accordance with observational studies, which did not reveal any effect of ACEi or ARB treatment on SARS-CoV-2 infection risk in individuals with hypertension or other CVDs^5^.

### ARB-treated COVID-19 patients have a reduced cell-intrinsic anti-viral response

We next assessed potential molecular mechanisms that may be involved in the delayed viral clearance of ARB-treated patients within the Pa-COVID-19 cohort described above. Pathway enrichment analysis based on the top 100 genes that were significantly differentially expressed (log fold-change > 0.25, FDR < 0.05, expression in > 10% of cells in one group) in either of the anti-hypertensive treatment groups compared to the HT-/CVD-group showed an activation of genes involved in stress and inflammatory response and antigen processing in ciliated cells of ARB+/HT+/CVD± COVID-19 patients (Figure 3a, Extended Data Figure 5a, Supplementary Table 4). For ACEi+/HT+/CVD± COVID-19 patients’ pathways related to defense response and regulation of viral genome replication were enriched in ciliated cells. Among those genes involved in regulation of viral genome replication, we found a number of statistically significantly upregulated type I interferon (IFN)-induced genes (e.g., *IFI6, IFI27, ISG15;* Figure 3a-b) in both treatment groups.

In secretory cells, ACEi treatment resulted in upregulation of genes negatively regulating immune system response to virus. Interestingly, ARB treatment led to a strong induction of genes involved in chemotaxis and inflammatory response in secretory cells (Figure 3a, Extended Data Figure 5a, Supplementary Table 4), including *CXCL1, CXCL6*, and *IL-8*, which recruit and activate neutrophils, and *CXCL17* which is a chemoattractant for monocytes, macrophages and dendritic cells (Figure 3a,c).

**Figure 3:**
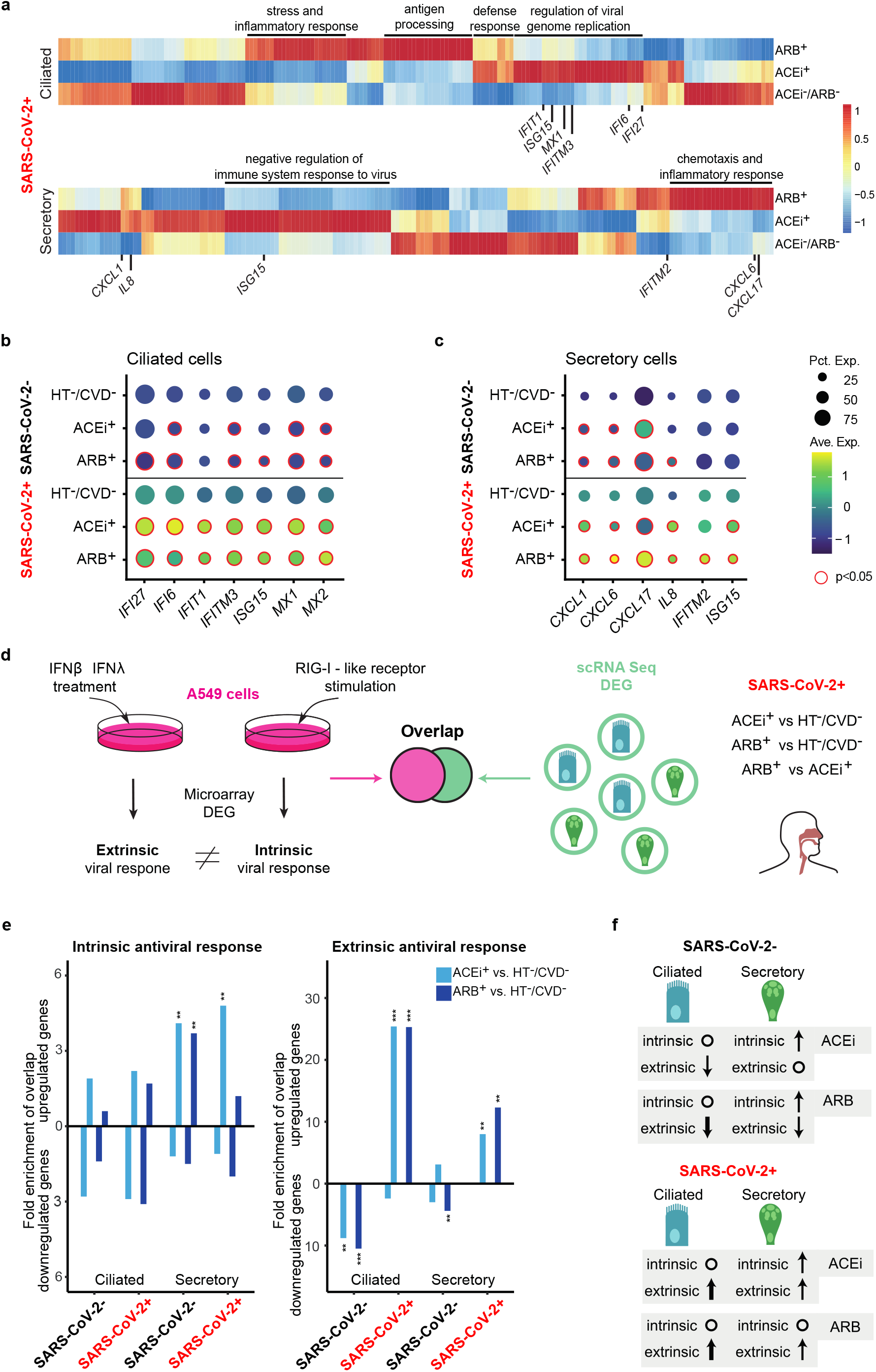
Differential regulation of antiviral response in patients with different anti-hypertensive treatments. (a) Scaled heatmaps showing the top 100 genes differentially expressed between SARS-CoV-2-positive ACEi+ or ARB+ and HT-/CVD-patients in ciliated and secretory cells by RNA sequencing (scaling by column). Enriched pathways (cf. Extended Data Figure 5a) and genes shown in b) are selectively marked next to heatmaps. (b,c) Expression plots of genes involved in regulation of viral genome replication in ciliated and secretory cells, respectively. Red circles indicate Benjamini-Hochberg adjusted two-tailed negative binominal *p*-value<0.05. Plotting labels on the right side. (d) Schematic layout of comparative overlap analysis of *in vitro* experiments (A549 lung cell culture) and single cell RNA sequencing of nasal swaps, displaying the workflow for generation of gene sets used in (e) and (f). For further details see Methods. (e) Bar plots showing the linear fold change of enrichment of overlap between the gene sets generated as shown in d) (intrinsic and extrinsic, left and right panel, respectively). The differentially regulated gene sets were split into up- and downregulated genes, which are displayed separately as positive and negative values on the x-axis, respectively. Asterisks indicate adjusted p-values derived from a hypergeometric test for overlap. ∗: p < 0.05, ∗∗: p < 0.01, ∗∗∗: p < 0.001. (f) Iconized table indicating the direction and strength of enrichment shown in e). Upward pointing arrows mark an enriched overlap in upregulated genes (ARB+ vs. HT-/CVD- and ACEi+ vs. HT-/CVD-), downward pointing arrows an enriched overlap in downregulated genes. Circles indicate that no significant enrichment of overlap is observed. The numbers of patients cohorts are, SARS-CoV-2-HT-/CVD-: n=6, SARS-CoV-2-ACEi+: n=6, SARS-CoV-2-ARB+: n=4, SARS-CoV-2+ HT-/CVD-: n=7; SARS-CoV-2+ ACEi+: n=10, SARS-CoV-2+ ARB+: n=15.

Next, we sought to disentangle cell-intrinsic responses triggered by viral infection and cell-extrinsic responses induced by signaling through type I/III IFNs. In an *in vitro* setting using A549 cells, we studied the extrinsic and intrinsic transcriptional response supposedly induced by SARS-CoV-2 infection. Cells were stimulated by either a highly specific RIG-I ligand triggering prototypical antiviral signaling through IRF3 or by a combination of IFNβ and INFλ inducing prototypical IFN signaling through ISGF3 (Figure 3d). Although the major pattern recognition receptor for SARS-CoV-2 remains elusive, all potential antiviral pathways converged on the transcription factors IRF3/IRF7 and NFkB^25^, eliciting a similar transcriptional response (Supplementary Table 4).

By overlapping the specific intrinsic and extrinsic antiviral response gene sets identified in the *in vitro* experiment with the differentially expressed genes in secretory and ciliated cells of COVID-19 patients (for enrichment see Methods), we observed that overall ACEi but not ARB treatment was associated with a strong cell intrinsic anti-viral response in SARS-CoV-2 positive patients (Figure 3e-f). Of note, already in SARS-CoV-2-negative patients, anti-hypertensive treatment by ACEi/ARB led to the induction of genes involved in the cell-intrinsic antiviral response in secretory but not in ciliated cells (Figure 3e-f). In secretory cells, pre-activation of the intrinsic antiviral response was further enhanced by ACEi-treatment of HT+/CVD± COVID-19 patients (Figure 3e). Surprisingly, intrinsic viral response was abolished in ARB+ treated HT+/CVD± COVID-19 patients. Extrinsic antiviral response genes were not pre-activated in SARS-CoV-2-negative patients treated by ACEi or ARBs. Upon SARS-CoV-2 infection, a robust extrinsic antiviral response was induced in both ciliated and secretory cells of HT+/CVD± COVID-19 patients treated by ACEi or ARBs (Figure 3e-f). A transcription factor binding motif analysis for genes differentially regulated in secretory cells confirmed the notion that the classical cell-intrinsic antiviral signaling through transcription factors such as IRF3, IRF1, and ISGF3 (ISRE) was enriched in ACEi+ treated HT+/CVD± COVID-19 patients (Extended Data Figure 5b). Instead, ARB+ treated patients showed a strong bias towards genes controlled by NFkB, a hallmark transcription factor for inflammatory conditions^26-28^.

Taken together, we observed a distinct difference in the balance between cell-intrinsic and extrinsic antiviral responses of ARB vs. ACEi treatment of HT+/CVD± COVID-19 patients. The identified dampened intrinsic antiviral response in secretory and ciliated cells of ARB-treated HT+/CVD± COVID-19 patients may have contributed to the above described delay in SARS-CoV-2 clearance in those patients.

### Crosstalk between epithelial and immune cells is associated with anti-hypertensive treatment in COVID-19 patients

The above described differential gene expression by ACEi/ARB revealed a distinct induction of inflammatory and chemoattractant genes. Hence, we inferred all possible intercellular interactions of all cell types and states across the different conditions using CellPhoneDB^29^ (Figure 4). Basal, secretory, ciliated, non-resident macrophages (nrMa), resident macrophages (rMa), neutrophils (Neu), and cytotoxic T cells (CTLs) had the highest number of interactions within the nasopharyngeal mucosa of COVID-19 patients (Figure 4a-b). A pre-existing cardiovascular comorbidity correlated with an increased number of cell-cell interactions with most of the previously mentioned cell types, gaining about 500 additional interactions upon SARS-CoV-2 infection (Figure 4a, Extended Data Figure 6a).

**Figure 4:**
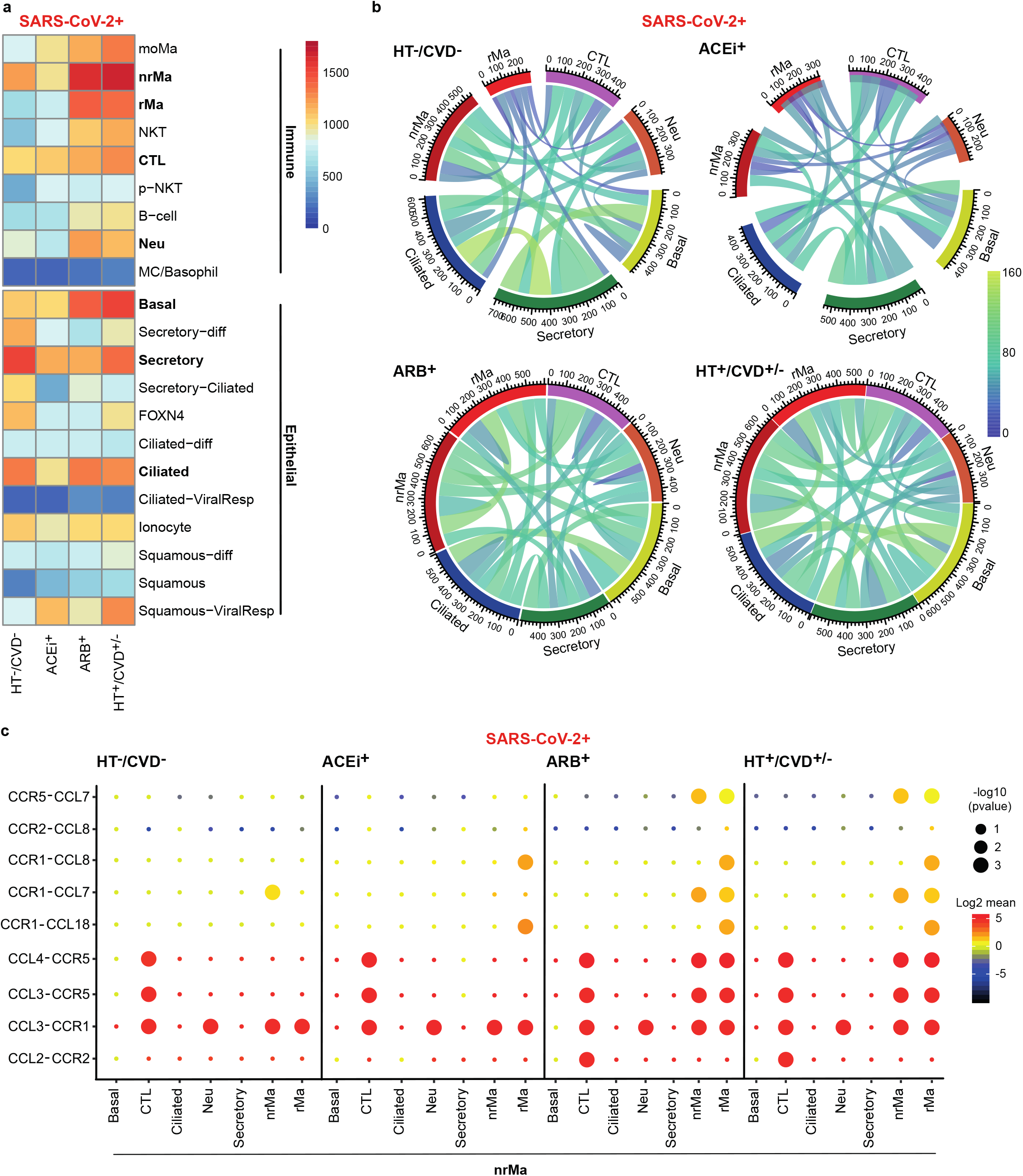
Cell-cell interactions in COVID-19 under anti-hypertensive treatment. (a) Heatmap depicting the total number of interactions per cell type across the different COVID-19 conditions. Scaled by number of identified interactions. (b) Circos plots of the most highly interactive cells (basal, secretory, ciliated, CTL, neu, nrMa, and rMa, printed in bold) scaled by the number of identified interactions. For circos plots of all cell types refer to Extended Data Figure 6a and 6b. (c) Dot plot showing immune modulatory interactions of nrMa across conditions with the highly interactive cells. Color-coding reflects log2mean expression while the p-value is shown by the dot size. The patient numbers for deriving the different sets were: SARS-CoV-2+ HT-/CVD-: 7; SARS-CoV-2+ ACEi+: 10; SARS-CoV-2+ ARB+: 15; SARS-CoV-2+ HT+/CVD±: 25.

In SARS-CoV-2 negative patients, interactions in ACEi+ and ARB+ were very similar in number and type (Extended Data Figure 6a-b). In contrast, for COVID-19 patients, ACEi treatment was concomitant with a reduction of interactions, while interactions in ARB treatment remained almost unchanged compared to HT+/CVD± patients.

The cell specific interactions were then categorized as intra- vs. inter-compartment interactions (immune:immune and epithelial:epithelial vs. immune:epithelial compartment interactions (Extended Data Figure 6c). In general, regardless of SARS-CoV-2 infection status, epithelial cells exhibited more potential interactions with themselves while immune cells had more inter-compartment interactions with epithelial cells. When comparing interactions in SARS-CoV-2-negative and -positive patients, we generally observed a loss of intra-compartment interactions for epithelial cells and a gain in inter-compartment interactions with immune cells among all conditions. Both inter- and intra-compartment interactions of immune cells tended to be increasen in HT+/CVD± compared to HT-/CVD-COVID-19 patients (Extended Data Figure 6c, Supplementary Table 5). Accordingly, intra-compartment interactions upon SARS-CoV-2 infection were exclusively statistically significantly increased in immune cell types, but decreased in epithelial cells (Supplementary Table 5).

Notably, this finding was mostly impacted by ARB-treated patients showing an overall increase in immune cell interactions, while ACEi-treated patients were similar to HT-/CVD-COVID-19 patients (Extended Data Figure 6c; Supplementary Table 5). In particular chemokine/chemokine receptor interactions mediated by nrMa (Figure 4c) reflected the similarity between HT-/CVD-patients and ACEi-treated COVID-19 patients. HT+/CVD+ and ARB-treated COVID-19 patients were similar in their interaction pattern, while in ACEi+ there was a reduced enrichment of interactions between *CCL3/CCL4* and *CCR5*, and between *CCR5* and *CCL7*, respectively (Figure 4c). In line with the pronounced chemokine/chemokine receptor interaction, the expression of *CCL2, CCL3, CCL4, CCL7*, and *CCL18* was upregulated in ARB+ concomitant with the expression of their receptors, e.g., *CCR1, CCR2*, and *CCR5*, suggesting a higher interactivity of nrMa under ARB compared to ACEi treatment (Extended Data Figure 6d).

### Hypertension-related inflammatory priming of immune cells is less pronounced in ACEi treated patients

To elucidate the deleterious contribution of hypertension on COVID-19, we evaluated the transcriptional profile of the key immune cell types orchestrating the antiviral response, namely T-cells, macrophages, and neutrophils. We and others showed that macrophages, in particular nrMa, are key mediators of hyperinflammation in severe COVID-19. High expression of genes coding for immune cell-recruiting chemokines, such as *CCL2, CCL3 and CCL4*, and inflammatory cytokines including *IL1B* and *IL8*, are hallmarks of nrMa in COVID-19^16, 17^. Upon SARS-CoV-2 infection, HT+/CVD± patients showed a significantly increased expression of these inflammatory mediators not only in macrophages but also in Neu compared to HT-/CVD-patients (Figure 5a, Extended Data Figure 7a). This hyperinflammatory phenotype was not only present in the upper airways but also in bronchial lavage (BL), as reflected by a stronger activation of BL-nrMa and BL-Neu of a hypertensive COVID-19 patient (BIH-SCV2-30) compared to a HT-/CVD-patient (BIH-SCV2-25, Extended Data Figure 7c). Anti-hypertensive treatment by ACEi apparently decreased hypertension-related hyperinflammation of COVID-19, while ARB was less effective in this regard (Figure 5a-c, Extended Data Figure 7b). In all macrophage subtypes, *CCL3* and *CCL4* expression among others was elevated in HT+/CVD±/ARB+ compared to HT+/ CVD±/ACEi+. In line, Neu showed a pro-inflammatory characteristic (*IL8, CXCL2)* and infiltrative potential (*ITGAM, ICAM1*) in ARB+ and to a much lesser extent in ACEi+, when compared to HT-/CVD-patients (Figure 5c).

**Figure 5:**
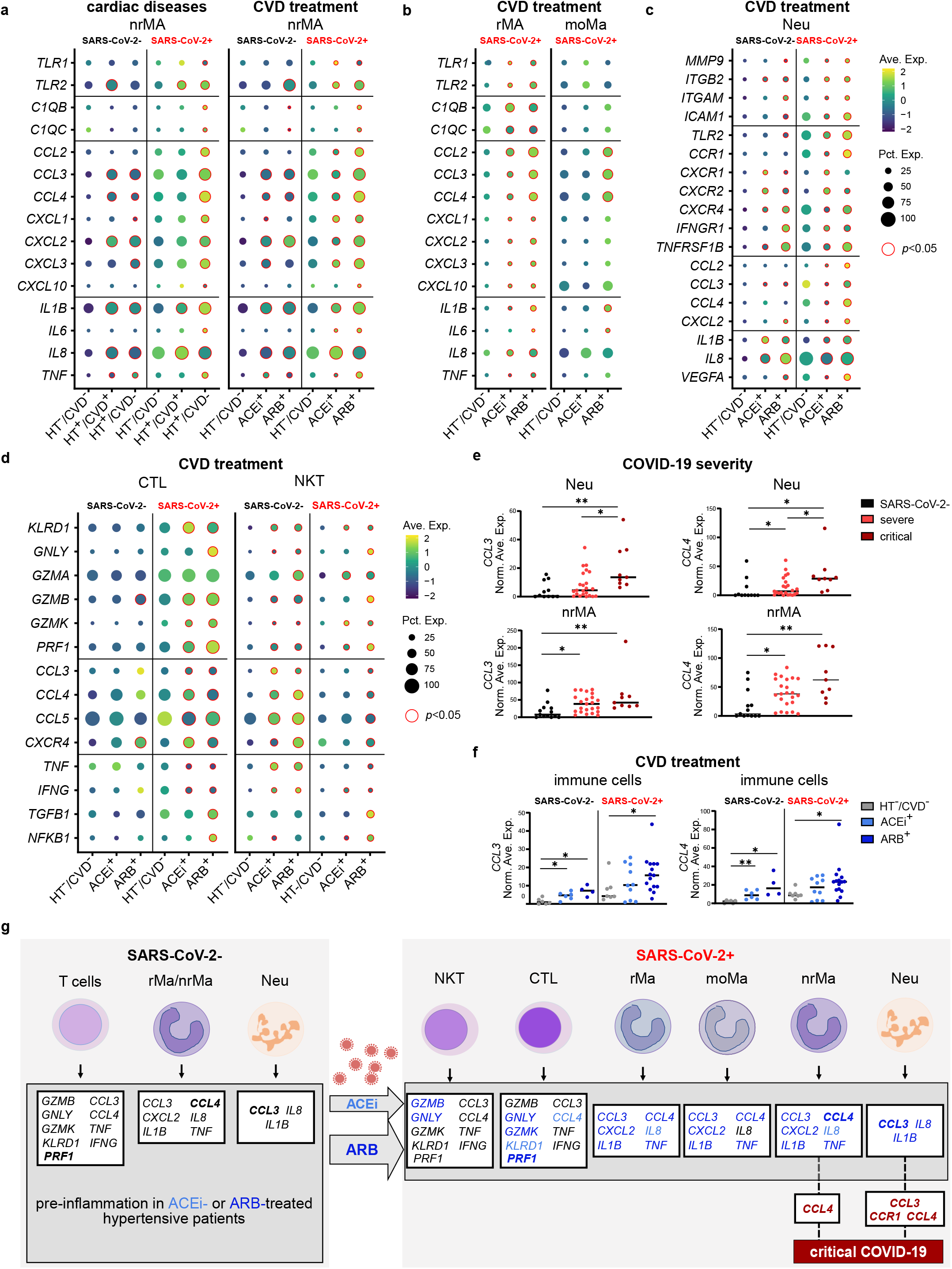
Hypertension-related immune response of the upper airway in COVID-19. (a-d) Dot plots depict gene expression of pro-inflammatory mediators, and receptors in macrophages, neutrophils, and T cells of the nasopharynx. Red circles indicate Benjamini-Hochberg adjusted two-tailed negative binominal *p*-value<0.05. Samples with no contributing cells per cell type were excluded from analysis. (a) Left panel depicts significant gene expression changes in nrMa of hypertensive patients with (HT+/CVD+, n (SARS-CoV-2-/+) = 4/10) or without an additional CVD (HT+/CVD-, n (SARS-CoV-2-/+) = 6/15) compared to HT-/CVD-patients (n (SARS-CoV-2-/+) = 6/6). (a-d) Significantly altered gene expression in rMa, moMa, Neu, NKT, and CTL of hypertensive patients treated either with ARB+ (n (SARS-CoV-2-/+) = 4/15), or ACEi+ (n (SARS-CoV-2-/+) = 6/10) in comparison to HT-/CVD-patients (n (SARS-CoV-2-/+) = 6/6). (e) Average gene expression of *CCL3* and *CCL4* in nrMa and Neu of SARS-CoV-2 negative (n=11 Neu, n=13 nrMa), severe COVID-19 (n=23), and critical COVID-19 patients (n=9). (f) Dot plots show average gene expression level of the MIP-1 encoding genes *CCL3* and *CCL4* across all immune cell types comparing ACEi+ and ARB+. (g) Left panel illustrates the immune cell priming of SARS-CoV-2 negative patients towards a pre-inflamed phenotype by HT independent of an ACEi or ARB treatment. Listed are all genes that are upregulated in SARS-CoV-2 negative patients with HT+, HT+/CVD+, ACEi+, and ARB+ in comparison to HT-/CVD-. As depicted in the right panel, upon infection, these genes are less activated during the anti-viral immune response against SARS-CoV-2 in ACEi+ (n=10) compared to ARB+ (n=15) treated patients. Shown are all pre-activated genes that are up-regulated in ARB+ (dark blue colored genes) and ACEi+ (light blue colored genes) compared to ACEi and ARB+, respectively. Black genes are not differentially expressed between ARB+ vs. ACEi+ treatment. The hypertension related exacerbated expression of *CCL3, CCL4*, and their receptor *CCR1* in Neu, and of *CCL4* in nrMa is associated with an increased risk for a critical clinical course of COVID-19 (genes depicted in dark red). Mann-Whitney U-test: ∗*p*-value<0.05, ∗∗*p*-value<0.005, ∗∗∗*p*-value<0.0005, Ave. Exp. = average gene expression, Pct. Exp. = percentage of cells expressing the gene.

Notably, in the absence of SARS-CoV-2 infection HT+/CVD- and HT+/CVD+ were characterized by inflammatory priming predominantly in nrMA and Neu (Figure 5a left panel, Figure 5c) - but not in rMa (Extended Data Figure 7b). In contrast to what we observed in COVID-19 patients, SARS-CoV-2-negative patients (HT+/CVD±/ACEi+ and HT+/ CVD±/ARB+) showed a similar chemokine fingerprint as reflected by an increased expression of e.g. *CCL3* and *CCL4* (Figure 5a right panel, Figure 5c).

In agreement with the previously observed aggravated cytotoxic capacity of CTLs in critical COVID-19 patients^16^, hypertensive patients treated with ARB or ACEi expressed cytotoxic mediators like *PRF1* or *GZMK* to a larger extend than HT-/CVD-patients upon infection with SARS-CoV-2 (Figure 5d). As expected, in the absence of SARS-CoV-2 infection, CTLs were not activated and no apparent difference between HT+/CVD±/ACEi+or HT+/CVD±/ARB+ and HT-/CVD-patients was observed (Figure 5d).

In contrast, in NKT cells cytotoxic markers (e.g. *KLRD1, GZMB*) in addition to monocyte-attractants (e.g. *CCL3, CCL4*) were already significantly elevated in SARS-CoV-2 negative hypertensive patients (Extended Data Figure 7a) independent of the type of anti-hypertensive treatment (Figure 5d). Upon infection, only in the ARB+ group apoptotic mediators (*e*.*g. GZMB*) and different chemokines (e.g. *CCL3, CCL4*) were significantly increased compared to HT-/CVD-.

Overall, we observed an increased expression of genes coding for pro-inflammatory and cytotoxic mediators in immune cells of hypertensive patients, already present before infection. ACEi but not ARB treatment apparently alleviated the hypertension-related inflammatory response to SARS-CoV-2 infection.

### Exacerbated expression of *CCL3* and *CCL4* observed in ARB-treated hypertensive patients correlates with disease severity

We next evaluated whether the hypertension-related inflammatory predisposition of nrMa, and Neu might contribute to an increased risk for critical COVID-19. *CCL3* and *CCL4* in nrMa and Neu, which were induced in HT+/CVD+/- compared to HT-/CVD- (Figure 5a-c), were statistically significantly elevated with the highest fold change in critical COVID-19 patients (n= 23 severe vs. 9 critical, cut-off: average fold change ≥0.25, and p-value≤0.05, Figure 5e). Using a logistic regression model considering age, gender, days post onset of symptoms and study center as potential confounding factors we confirmed a significant relationship between increased expression of *CCL4* derived from either nrMa (adj.OR/95% CI=1.04/1.00-1.07, p-value=0.027) or Neu (adj.OR/95% CI=1.06/1.00-1.12, p-value=0.044) and *CCL3* expressed by Neu (adj.OR/95% CI=1.13/1.01-1.27, p-value=0.02) and an increased risk for critical COVID-19. Notably, expression of *CCR1*, the receptor bound by CCL3 and CCL4, increased in nrMa and Neu with COVID-19 severity supporting the potential of CCR1 as a therapeutic target^16^ (Extended Data Figure 7d). Antihypertensive treatment increased *CCL3* and *CCL4* expression in immune cells of SARS-CoV-2 negative patients, but in COVID-19 only ARB-treatment significantly elevated *CCL3*/*CCL4* expression compared to HT-/CVD-COVID-19 (Figure 5f).

In summary, we could show that treatment with ACEi resulted in a more favorable immune response after SARS-CoV-2 infection, while ARB therapy did not sufficiently alleviate the hypertension-related hyperinflammation especially in nrMa and Neu, possibly contributing to critical COVID-19 course (Figure 5g).

## Discussion

This study identified potential novel molecular mechanisms underlying the finding from observational studies that COVID-19 patients with hypertension or coronary artery disease revealed higher morbidity and mortality rates^4, 30, 31^. As first line anti-hypertensive medication includes modulators of RAAS interfering with the pathway employed by SARS-CoV-2 for cellular entry it has been debated whether ACEi or ARB treatment alters SARS-CoV-2 infectivity and severity of COVID-19. Our data suggest that the hypertension-associated additional risk for critical disease progression can be reduced by ARB treatment and is almost abolished by ACEi treatment. This is corroborated by previous reports observing higher mortality rates in hypertensive COVID-19 patients in the absence of ACEi/ARB treatment^32^.

Several clinical studies are now available comparing SARS-CoV-2 infectivity rates among patients with and without ACEi/ARB treatment^13, 33^. Their findings support the notion that testing positive for SARS-CoV-2 is not associated with treatment by ACEi/ARB^13, 34^. In line, we observe no difference in *ACE2* expression and initial viral concentration between patient groups. Also, induction of ACE2 expression after SARS-CoV-2-infection was not altered by ACEi/ARB treatment. However, viral clearance was delayed by ARB treatment. While reduced viral clearance can be a result of defects in immunity for example of an impaired T cell activity, as it has already been reported for cardiovascular diseases^35^, our data suggest that the altered anti-viral-response of ciliated and secretory epithelial cells plays a crucial role. Intrinsic anti-viral response was dampened in ARB-treated patients, while patients under ACEi treatment showed similar viral clearance as normotensive patients together with an elevated intrinsic antiviral response.

We identified hypertension-associated elevated immunological activity as the prominent factor contributing to the increased risk of hypertensive patients for COVID-19 severity. Hypertensive patients showed an inflammatory predisposition of different immune cell subtypes observed already before SARS-CoV-2 infection irrespective of anti-hypertensive treatment. Upon SARS-CoV-2 infection, ARB-treated patients showed an exaggerated hyperinflammatory response, which was alleviated in ACEi treated patients. This distinct inflammatory response of COVID-19 patients under different anti-hypertensive treatment may also give rise to the here described less pronounced risk reduction for disease severity under ARB compared to ACEi therapy.

Interestingly, a recent study showed enhanced plasma ACE2-activity along with a significant increase in Ang(1-7) concentrations in ACEi treated COVID-19 patients compared to COVID-19 patients under ARB therapy, suggesting a higher anti-inflammatory capacity in ACEi compared to ARB treated COVID-19^36^.

The present study demonstrated an immune activation in hypertensive patients that is largely augmented under COVID-19 and may provide a novel explanation for the adverse course of the disease in these patients related to a hyperinflammatory response. Our data are in line with the general guideline recommendations discouraging discontinuation of ACEi or ARB treatment. On the contrary, our results may suggest that ACEi could be the more beneficial antihypertensive treatment during COVID-19. A randomized control trial is required to assess the clinical impact of ACEi vs. ARB treatment in COVID-19 patients and several trials are under way.

## Data Availability

Due to potential risk of de-identification of pseudonymized RNA sequencing data the raw data will be available under controlled access in the EGA repository, [will be added upon manuscript acceptance]. Count and metadata tables (patient-ID, sex, age, cell type, QC metrics per cell) can be found at FigShare: [will be added upon manuscript acceptance]. In addition, these data can be further visualized and analyzed in the Magellan COVID-19 data explorer at https://digital.bihealth.org [will be publicly available upon manuscript acceptance].

## Methods

### Patient Recruitment and Ethics Approval

Patients were enrolled between March 6^th^ and June 7^th^ 2020 in either the prospective observational cohort study Pa-COVID-19^21^ at Charité – Universitätsmedizin Berlin or the SC2-study at the University Hospital Leipzig. Written informed consent was given by all patients or their legal representatives. The study was approved by the respective Institutional Review boards of the Charité-Universitätsmedizin Berlin (EA2/066/20) or the University Hospital Leipzig (123/20-ek) and conducted in accordance with the Declaration of Helsinki.

### Pa-COVID-19 cohort

Between March–May 2020, 162 COVID-19-positive patients were recruited at Charité – Universitätsmedizin Berlin in the Pa-COVID-19 study. In the here presented study, we excluded those patients who had their positive SARS-CoV-2 test exclusively outside the Charité (n=12) and those with missing information on ACEi/ARB treatment (n=6). For the remaining 144 COVID-19 patients, we assessed differences in COVID-19 severity related to pre-existing cardiovascular diseases (CVD+), such as hypertension (HT+/CVD-) or HT and an additional cardiovascular disease (coronary artery diseaseand/or heart failure, HT+/CVD+) in the different treatment groups (ACEi+, ARB+, ACEi-/ARB-) compared to patients without HT-/CVD-. HT was defined according to ESC/ESH guidelines as office blood pressure ≥ 140mm Hg systolic or ≥ 90 mm Hg diastolic^37^. The characteristics of this cohort are summarized in Supplementary Table 1 and 2.

### Single-cell RNA sequencing cohort

We collected nasopharyngeal swabs for single-cell RNA transcriptome analyses (scRNA-seq) of 32 confirmed COVID-19 patients (23 males and nine females; median age: 67; range: 32–91 years of age) and 16 controls without any COVID-19 related symptoms (nine males, seven females; median age: 53; range: 24-79). Relevant patient characteristics are given in Supplementary Table 3.

Of the COVID-19 patients, 23 patients were classified as having severe, and nine as having critical disease according to the World Health Organization (WHO) guidelines^38^. Hospital mortality of these patients was 1/33 (3.0%).

Baseline CVD were prevalent in 25 of the COVID-19 patients. They either suffered from HT only (n=16, 64%), CVD with HT (n=7, 28%), or CVD with heart failure and HT (n=2, 8%). Patients with HT only were classified as HT+, while all patients suffering from HT and CVD (and possibly additionally heart failure) were classified as HT+/CVD+. Concomitant treatment of HT+/CVD± included treatment with either ACEi (n=10) or ARB (n=15). Seven of the COVID-19 patients had no known CVD (21.2%). Note that patient BIH-SCV2-14 was included in all analysis regarding ACEi/ARB treatment but not in the CVD analysis (this patient suffered from HT and heart failure). Of the symptom-free SARS-CoV-2-negative patients, six had HT only (37.5%), and four had additional CVD (25%). Six of these patients received ACEi (37.5%) and four were treated with ARB (25%). The remaining six patients had no known CVD and therefore, were neither treated with ACEi nor ARB (37.5%).

### Isolation and preparation of single cells from human airway specimens, followed by pre-processing of the raw sequencing reads

Sample procurement, single-cell isolation, library preparation, and subsequent data analysis was performed as described previously^16^. Briefly, freshly taken nasopharyngeal swabs from donors were directly transferred into 500 μL cold

DMEM/F12 medium (Gibco, 11039) and 500 μL of 13mM DTT (AppliChem, A2948) were added to each sample. Cells were released by gently pipetting the solution onto the swab, followed by dipping the swab 20 times into the medium. Subsequently, the samples were incubated on a thermomixer at 37°C, 500 rpm for 10 minutes, followed by centrifugation at 350xG at 4°C for 5 minutes. While carefully removing the supernatant, the pellet was visually examined for any traces of blood. If it contained red blood cells (RBC), the pellet was resuspended in 500 μL 1x PBS (Sigma-Aldrich, D8537) and 1 mL of RBC Lysis Buffer (Roche, 11814389001), incubated at 25°C for 10 minutes, and subsequently centrifuged at 350xG at 4°C for 5 minutes. A single cell suspension was achieved by resuspending the cell pellet in 500 μL Accutase (Thermo Fisher, 00-4555-56), followed by incubation at room temperature for 10 minutes with gently mixing the cells after 5 minutes by pipetting. Subsequently, 500 μL DMEM/F12 supplemented with 10% FBS was added to the cells. After centrifugation at 350xG at 4°C for 5 minutes and removal of the supernatant, the cell pellet was resuspended in 100-500 μL 1x PBS (depending on the size of the cell pellet). Cell debris was removed by using a 35 μm cell strainer (Falcon, 352235) before cell counting was performed using a disposable Neubauer chamber (NanoEnTek, DHC-N01). The cell suspension was loaded into the 10x Chromium Controller using the 10x Genomics Single Cell 3’ Library Kit v3.0 (10x Genomics; PN 1000076; PN 1000077; PN 1000078) and 10x Genomics Single Cell 3’ Library Kit v3.1 (10x Genomics; PN 1000223; PN 1000157; PN 1000213; PN 1000122) and the subsequent reverse transcription, cDNA amplification, and library preparation was performed according to the manufacturer’s instructions. Importantly, we extended the incubation at 85°C during the reverse transcription to 10 minutes to ensure virus inactivation. Afterwards, the 3’RNA sequencing libraries were pooled either for S2 or S4 flow cells (S2: up to eight samples, S4: up to 20 samples) and sequenced on the NovaSeq 6000 Sequencing System (Illumina, paired-end, single-indexing).

All samples were processed under biosafety S3 within one hour after procurement. Note that samples not immediately used for library preparation were resuspended in cryopreservation medium [20% FBS (Gibco, 10500), 10% DMSO (Sigma-Aldrich, D8418), 70% DMEM/F12] and stored at −80°C. Frozen cells were thawed quickly at 37°C, pelleted at 350xG at 4°C for 5 minutes, and proceeded with normal processing.

Single-cell datasets were processed using cellranger 3.0.1. All transcripts were aligned to a customized human hg19 reference genome (10x genomics, version 3.1.0) plus the SARS-CoV-2 genome (Refseq-ID: NC_045512) added as an additional chromosome. Following alignment, ambient RNA was removed using SoupX^39^ using *MUC1, MUC5AC*, and *MUC5B* as marker genes. Where ambient RNA levels seemed plausible (5-15%), filtered expression matrices were used for downstream analyses. Further processing was performed using Seurat 3.1.4. Genes were retained if they were present in at least three cells in a sample. Cells with more than or equal to 15% mitochondrial reads or less than 200 genes expressed were removed from the analysis. For the number of UMIs, an upper cutoff was chosen manually per sample based on outliers in a UMI counts vs. gene counts plot and was typically in the range of 75,000 to 150,000. After normalizing to 10,000 reads per cell, samples were integrated using stepwise CCA on smaller subsets using 90 components and 2,000 variable genes identified by SelectIntegrationFeatures. On the integrated dataset, PCA was run using 90 principal components, followed by UMAP and clustering with a resolution of 2.1, both using all components. NKT, CTL, and p-NKT cells were subsetted for further analysis. Scaling, dimensional reduction by PCA and the UMAP was calculated separately for this subset.

Cell types were then refined manually by assessing the expression of known cell type markers. Cell types from epithelial^22, 40, 41^ and immune^42^ cell populations were identified according to the expression levels of different marker genes (Extended Data Figure 1c). The “viral responsive” cell states of ciliated and squamous epithelial cells (Extended Data Figure 1b) were identified by gene set enrichment analysis using clusterProfiler version 3.12.0 and the output of “FindClusters()” function from Seurat as input (https://yulab-smu.github.io/clusterProfiler-book/index.html)^43^.

For cell-cell interactions, which are based on the expression of known ligand-receptor pairs in different identified cell types, CellPhoneDB^21^ version 2.1.2 was used (https://github.com/Teichlab/cellphonedb). circlize 0.4.10 was used to generate the circos plots to display the cell-cell interactions^44^.

Shifts of interactions across the different conditions were tested for significance using a logistic regression based on a binomial distribution (Supplementary Table 5). Arboreto^45^ 0.1.5 and pySCENIC^46^ 0.10.0 were used to infer transcription factor importance.

### Viral load measurement

SARS-CoV-2 RT-PCR results and SARS-CoV-2 RNA concentrations were obtained by using respiratory samples taken for routine testing and by using two different test systems. First, by using an assay targeting the SARS-CoV-2 E-gene as published before^47^ and the Roche LC480 instrument. Second, by using the cobas® SARS-CoV-2 test on the cobas® 6800/ 8800 system. In cases of the LC480 system, RNA was extracted by using the MagNA Pure 96 DNA and Viral NA Small Volume Kit on a Roche MagNA Pure 96 system. We quantified SARS-CoV-2 RNA by applying external calibration curves and quantified *in vitro* transcribed RNA, derived from the E-gene fragment^47^ or purified complete SARS-CoV-2 RNA. Viral load (using the E-gene genome target for both test systems) was calculated taking into account different predilutions, extraction volumes, and RT-PCR reaction volumes.

### Viral load assessment by regression analysis

Data on positive viral mRNA measurements were available for 144 patients of the Pa-COVID-19 cohort. To smooth the longitudinal viral mRNA data of each patient, the values were binned in three-day intervals with respect to the time of the first test result. The maximal value in each bin was considered. In case a patient had a negative test for SARS-CoV-2 between two positive measurements, the negative result was disregarded. Patients for which only one negative test result was available or had missing confounder information were excluded from the analysis. A linear repeated measurement mixed model assuming a heterogeneous first-order autoregressive structure of the covariance matrix was applied considering ACEi or ARB treatment in comparison to untreated patients without pre-existing CVD - otherwise treated HT+/CVD+/- patients were excluded - as a fixed effect. The concentration measurements up to the fifths consecutive viral test were included in the model that was adjusted for days post onset of symptoms, gender, BMI, smoking, and insulin treatment. Calculations were performed in SPSS version 25 and predicted means calculated using the maximum likelihood option.

### Slope analysis of viral clearance

To compare the rate of viral clearance between patient groups, we performed a linear fit to viral load measurements during the first 30 days post symptom onset. Zero measurements, and patients with fewer than four non-zero measurements within this timeframe, were excluded from this analysis. Student’s t-tests were used to identify statistically significant differences in slope between groups.

### Identification of RIG-I and type I/III interferon responsive gene sets

A549 cells were electrotransfected with 400bp long in vitro transcribed dsRNA^48^ and lysed at 2, 4, 6, 8, 16 and 24h after transfection, or mock electrotransfected and lysed at 2 and 24h. Alternatively, A549 cells were treated with a mix of 100 IU/ml interferon beta (8499-IF-010/CF, R&D Systems, Minneapolis, MN, USA) and 2.5 ng/ml interferon lamba-1 (300-02L-100, Peprotech, Hamburg, Germany) for 2, 8 or 24h and then lysed. Total RNA was extracted from cell lysates using the NucleoSpin RNA Plus kit (Macherey Nagel, Düren, Germany) and the RNA was subjected to microarray analyses using the Illumina Human HT-12 Expression Beadchip platform at the genomic and proteomics core facility at DKFZ. Expression data was quantile normalized, genes with no significant expression at any condition / time point were excluded and gene regulation at different treatment time points vs. the 0h control was determined using the limma package (Bioconductor). Data were then filtered according to the following criteria to define gene sets.

The gene set “Cell-intrinsic antiviral (RIG-I-like receptor, RLR) signaling” comprises genes that were exclusively or predominantly upregulated upon dsRNA transfection (RLR stimulation) but not upon IFN treatment; while for the sake of specificity a very specific RIG-I stimulation was applied, the transcriptional response likely is similar for any antiviral stimulus (e.g. through MDA5, STING, TLRs) that activates the IRF3 transcription factor. The Cut-offs were as follows: maximum (at any time point) log2-fold change in dsRNA transfected samples (maxLog2FC-RNA) > 2.0 and maximum log2-fold change in IFN treated samples (maxLog2FC-IFN) <1.0; genes were excluded as electrotransfection artifacts if maximum log2-fold change in mock transfection (maxLog2FC-mock) > 0.5∗(maxLog2FC-RNA – maxLog2FC-IFN) (11 genes). This procedure yielded a list of 238 genes, comprising expected genes such as the IRF3-dependent type I and III IFN genes themselves (*IFNB1, IFNL1,2,3*) and classical NFkB targets such as *TNFAIP3* (previously *A20*) and the IkB genes *NFKBIA, NFKBIB, NFKBIZ*.

The gene set “Extrinsic / paracrine type I / III IFN signaling” comprises genes that were strongly upregulated by extrinsic IFN beta / IFN lambda treatment while less so by cell-intrinsic RLR induction. Note that the majority of IFN-induced genes were upregulated by RLR signaling as well, putatively due to the above noted IFN production upon RLR stimulation. We enriched this gene set for genes with a bias towards IFN and against RLR signaling by applying the below described filters. Notably, a few genes, such as *LY6E*, described to possess antiviral activity against SARS-CoV-2^49^, were induced only upon IFN treatment but not at all by RLR signaling. In general, we found less profound gene induction in IFN-treated than in dsRNA transfected conditions, likely due to the moderate dose of IFN used; we therefore used less stringent cut-offs for this gene set: maxLog2FC-IFN > 0.8 and (maxLog2FC-IFN - maxLog2FC-RNA) > −0.5. The latter filter removed roughly 50% of the genes, selecting for those with a relative bias of IFN-treatment over dsRNA transfection. Filtering yielded 95 genes, including many of the well-known *ISGF3*-driven IFN-stimulated genes (ISGs), including the MX-family genes, *IFIT1, IFITM2*/*3* and *ISG15*.

### Statistics

Differences in the percentage of *ACE2* expressing cells were calculated using logistic regression in R 3.5.1. 95% confidence intervals for the percentage of *ACE2* expressing cells are provided. Differences in gene expression were calculated using “FindMarkers()” in Seurat version 3.1.4 with the MAST-based differential expression test adjusted for days post symptoms onset (dps). Overlap statistics were calculated as hypergeometric tail probabilities. Differences in CPM were calculated using an ANOVA followed by Tukey’s honest significance of differences test after assessing homoskedasticity using Bartlett’s test. When multiple tests were performed, p-values were adjusted using the Benjamini-Hochberg method. Total CPM values were extracted from the filtered and raw matrices output by CellRanger. Motif enrichment p-values were calculated using HOMER 4.10.0^50^. To analyze the potential contribution of HT/CVD and its treatment on COVID-19 severity in the PaCOVID-19 cohort we conducted chi-square tests with Yates’ correction and logistic regression models adjusted for gender, BMI smoking and insulin treatment. Age showed collinearity with ACEi or ARB treatment. Therefore, age was omitted as a confounder in models where ACEi or ARB treatment was used as an independent variable. Viral clearance was assessed based on similarly adjusted linear regression models as described above. Logistic regression models assessing gene expression changes observed in the scRNA-seq cohort were adjusted for age, gender, days post onset of symptoms and study center.

### Code availability

No custom code was generated/ used during the current study.

## Acknowledgement

We thank all patients of the Pa-COVID-19 cohort study for kindly donating nasopharyngeal samples and clinical data. We also thank Alexander Krannich and Julia Kazmierski, Charité – Universitätsmedizin Berlin, for help in sample procurement/annotation and for supporting sample processing, respectively. We thank Antje Seidel and Grit Szczepankiewicz for supporting patient recruitment at the University Hospital Leipzig. We thank Carola Spann and Darius Schweinoch for their help with the *in vitro* experiment. This study was supported by the BIH COVID-19 research program, the European commission (ESPACE, 874719, Horizon 2020), the BMBF-funded de.NBI Cloud within the German Network for Bioinformatics Infrastructure (de.NBI; 031A537B, 031A533A, 031A538A, 031A533B, 031A535A, 031A537C, 031A534A, 031A532B), and the BMBF-funded Medical Informatics Initiative (HiGHmed, 01ZZ1802A - 01ZZ1802Z). We thank Illumina GmbH for financial support via the allocation of reagents and sequencing flow cells as well as Markus Vossmann, Martin Allgaier, Oliver Krätke for the realization of the sequencing runs at the Illumina Solutions Center Berlin. MSA has received research support from the German Cardiovascular Research Center.

## Authors contributions

R.E., C.C., U.L., I.L. conceived, designed, and supervised the project. V.C. and C.D. performed qPCR experiments and/or provided the data, Sa.T., S.L., R.L.C. L.T., T.K., C.K. performed data analysis. Jo.L., R.L.C., Je.L., M.M. performed the single-cell RNA sequencing experiments. M.B. performed and analyzed the *in vitro* experiment. Je.L., C.K., M.M., J.K., F.P. provided experimental support. M.S.A, S.H., A.L., B.H., F.K., M.W., M.T.V., S.D.M, U.G.L., L.-E.S. and Sv.L. provided the human specimens, clinical data and annotation of the patients. Sa.T. and B.P.H. managed patient data of the cohort. Sv.T., J-P.A., J.E. provided technical and data management support. Sv.T. developed Magellan. B.P.H., C.G., N.I., L.K. and U.L. contributed with discussion of the results. Sa.T., Sv.L., S.L., R.L.C., L.T., B.P.H., R.E. and I.L. wrote and prepared the manuscript. All authors read, revised, and approved the manuscript.

## Main Figures

**Extended Data Figure 1:**
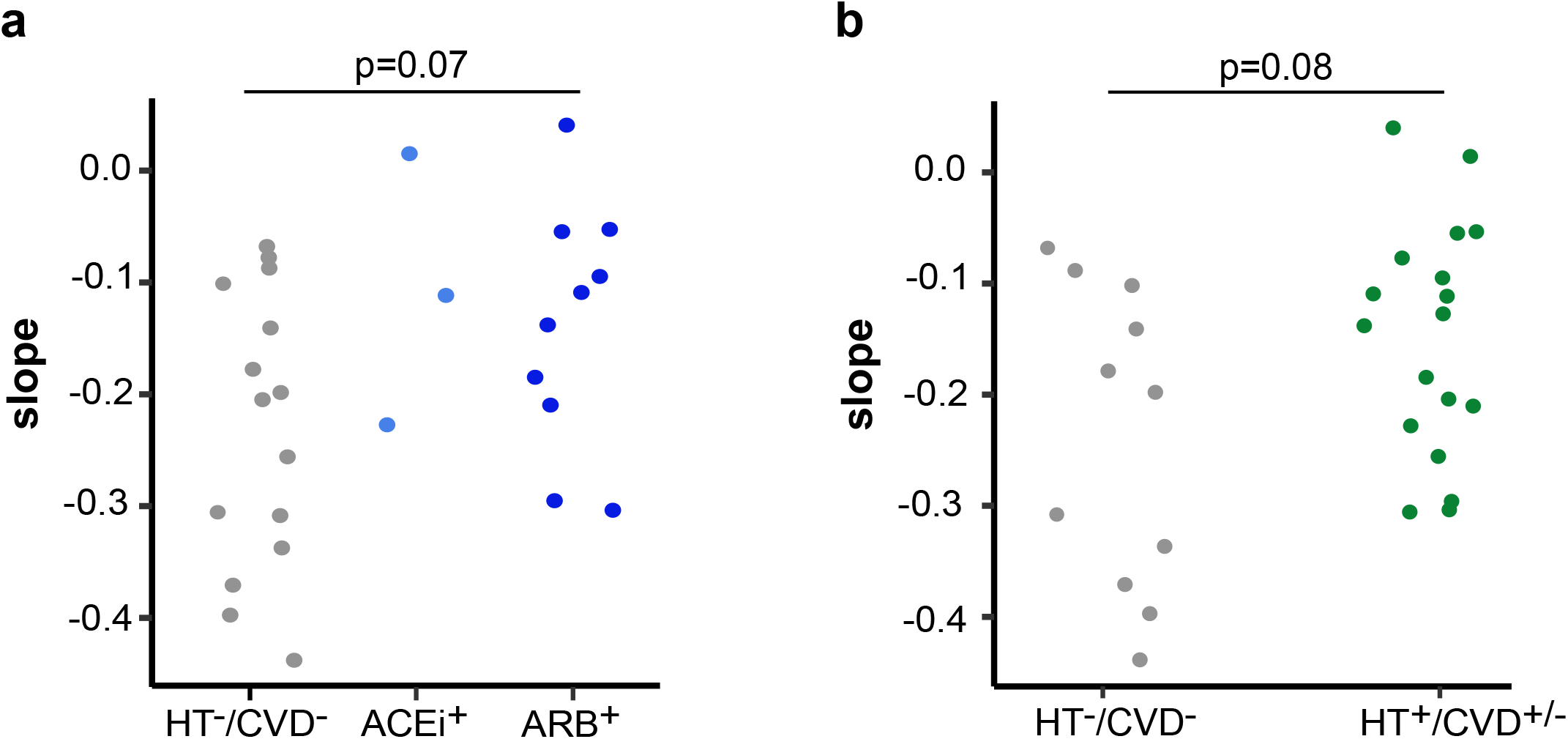
Slope analyses of viral clearance for Pa-COVID-19 patients. Slope analyses (a) of the viral clearance related to ACEi or ARB treatment and (b) in in relation to hypertension. CVD = cardiovascular disease, HT = hypertension, p-values (p) from Student’s t-test.

**Extended Data Figure 2:**
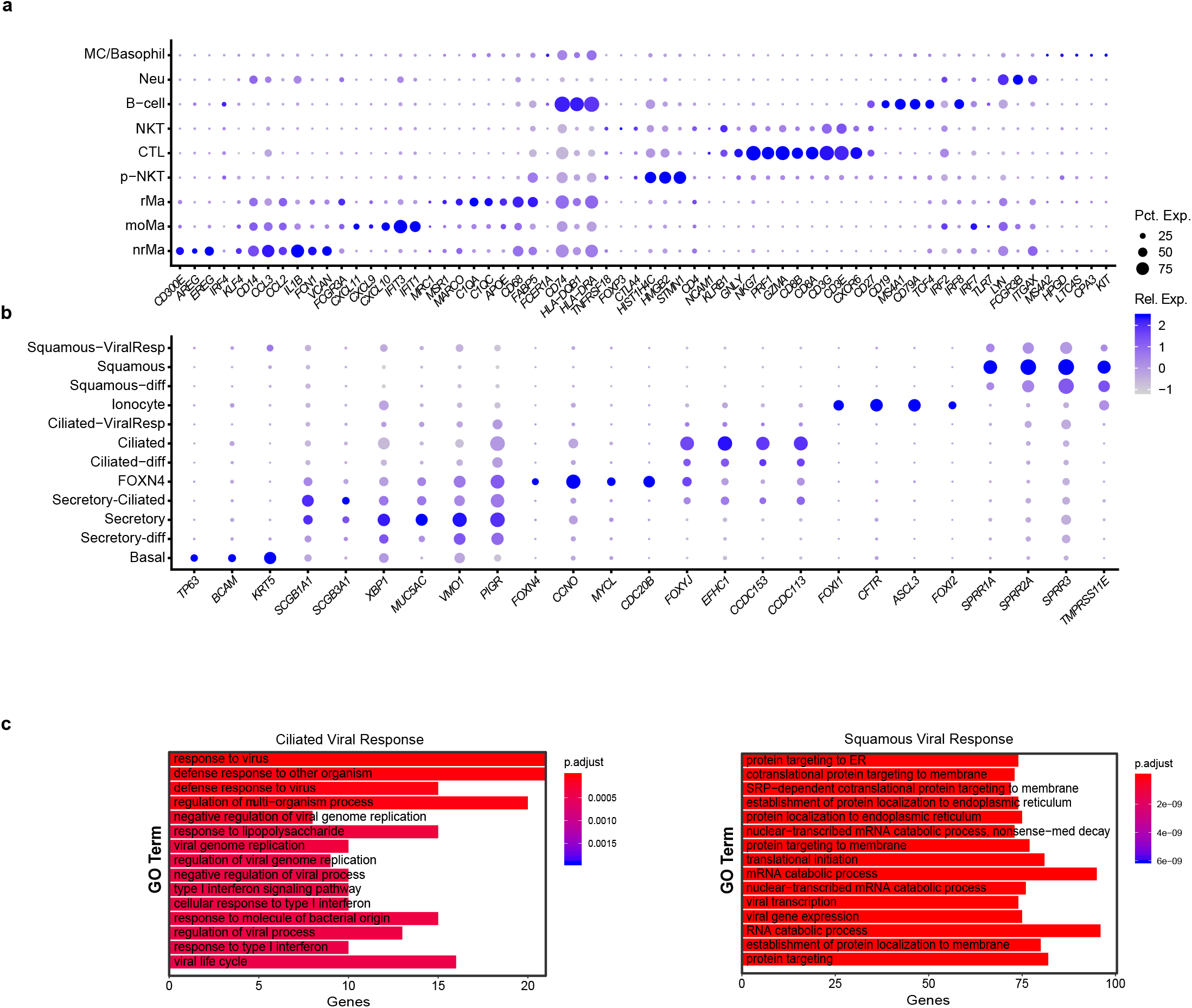
Cell type characteristics of the scRNAseq cohort. (a) Dotplot depicting marker genes used to identify immune cell types. (b) Dotplot depicting marker genes of epithelial cell types. For dotplots, expression levels are color-coded and the percentage of cells expressing a respective gene is size-coded. (c) GO-term enrichment for differentially expressed genes in the ciliated and squamous ViralResp cell cluster. Number of genes observed per term are depicted, colour indicates adjusted p-values.

**Extended Data Figure 3:**
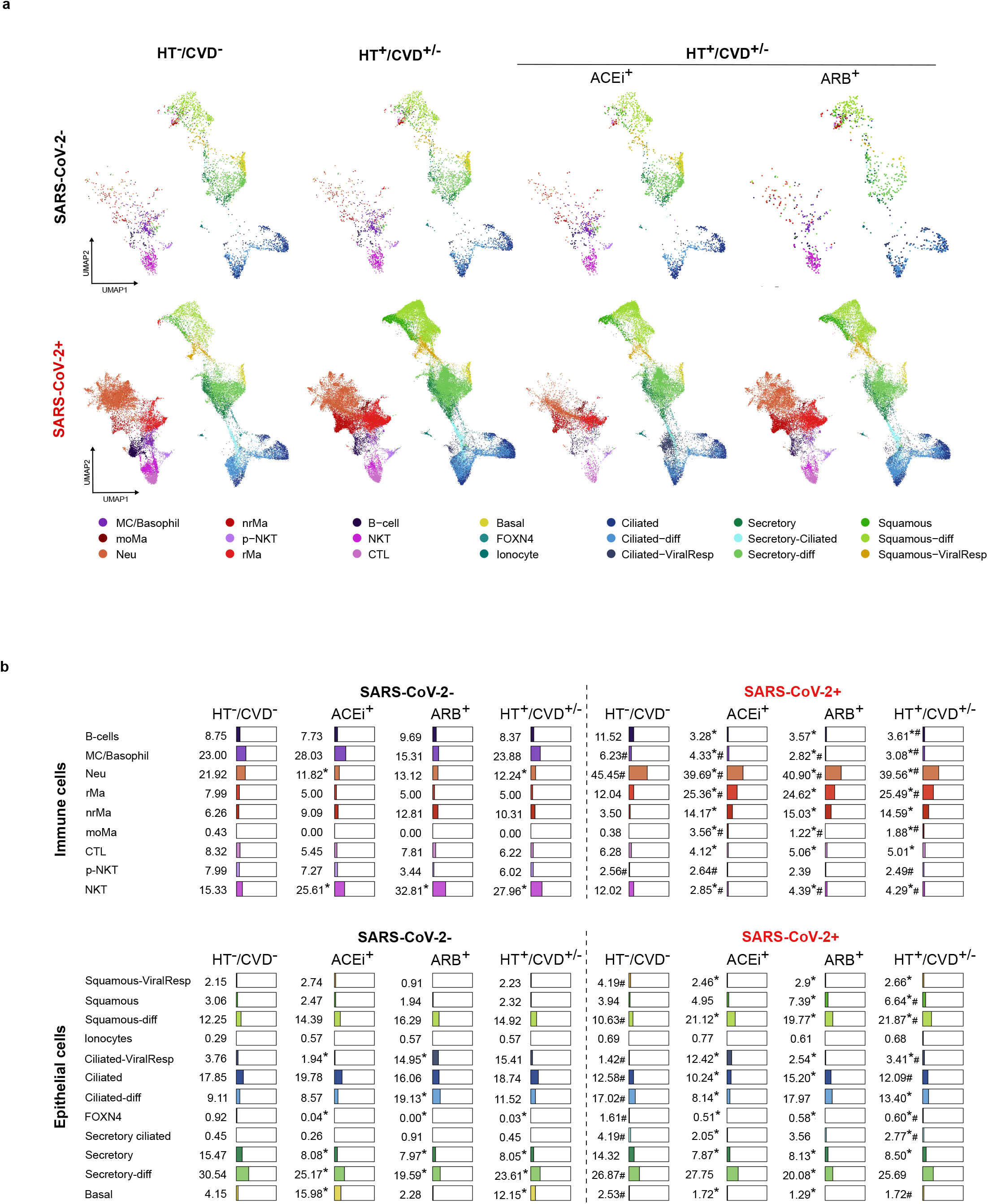
Cell type distribution in the scRNA-seq cohort. (a) UMAPs depicting distribution of cell types and states in SARS-CoV-2 negative and positive patients. UMAPs are presented separately for controls (HT-/CVD-), patients with a pre-existing cardiac disease (HT+/CVD±) and the treatment thereof (ACEi+ and ARB+, respectively). Pct. Exp. = percentage of cells expressing the gene; Rel. Exp. = relative gene expression. (b) Distribution of immune and epithelial cell types/states in SARS-CoV-2 negative and positive patients separated by HT-/CVD-, HT+/CVD± or ARB/ACEi-treatment. Given are percentages related to the total number of immune or epithelial cells respectively. ∗sign. compared to HT-/CVD-, #sign. compared to SARS-CoV-2 neg. UMAP = uniform manifold approximation and projection, MC = mast cells; moMa = monocyte derived macrophage; Neu = neutrophil; rMa = (non-)resident macrophage; p-NKT = proliferating natural killer T-cell; CTL = cytotoxic T lymphocyte; diff = differentiating; ViralResp = viral response.

**Extended Data Figure 4:**
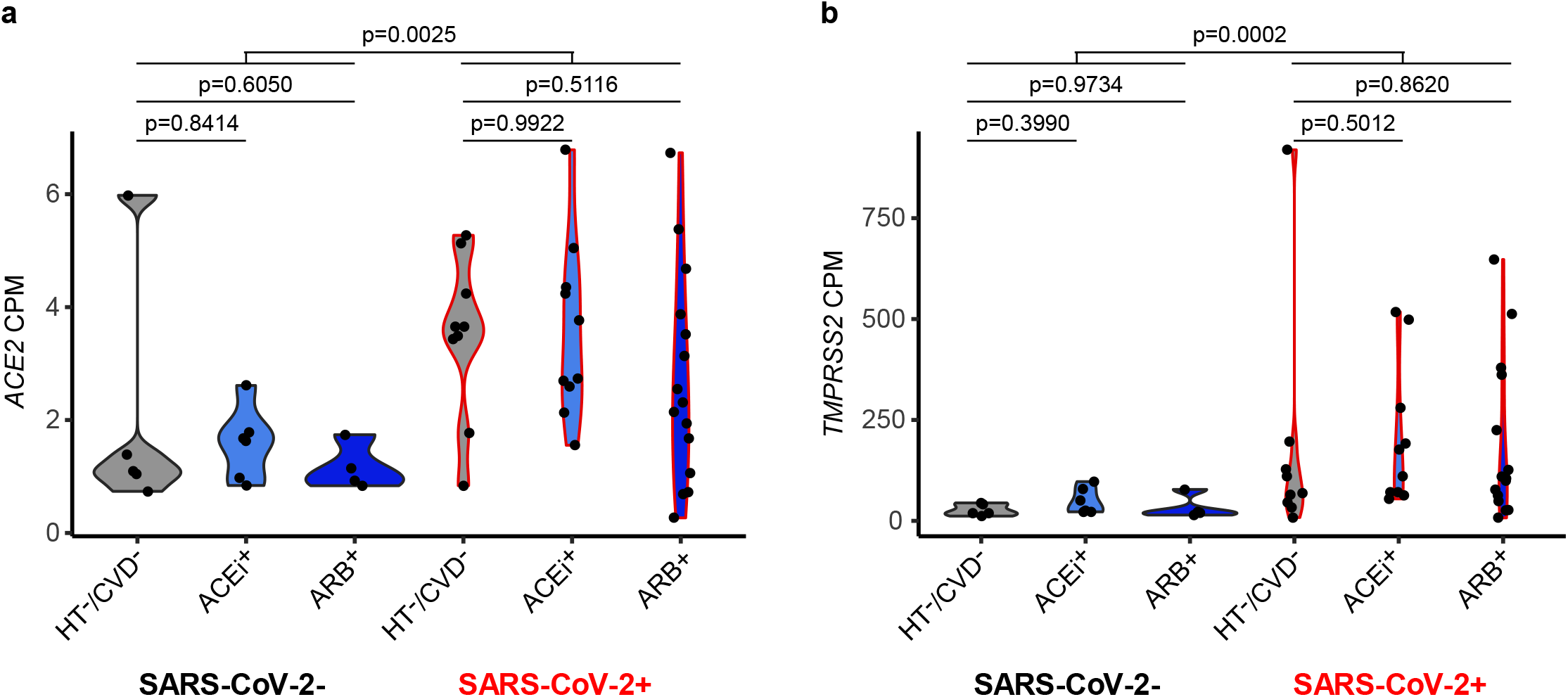
*ACE2* and *TMPRSS2* expression. Violin plots showing the total *ACE2* and *TMPRSS2* expression per sample in counts per million, split by treatment and infection group. Tukey’s p-values for individual comparisons are shown. The patient numbers for the different sets were: SARS-CoV-2-HT-/CVD-: 6; SARS-CoV-2-ACEi+: 6; SARS-CoV-2-ARB+: 4; SARS-CoV-2+ HT-/CVD-: 8; SARS-CoV-2+ ACEi+: 10; SARS-CoV-2+ ARB+: 15.

**Extended Data Figure 5:**
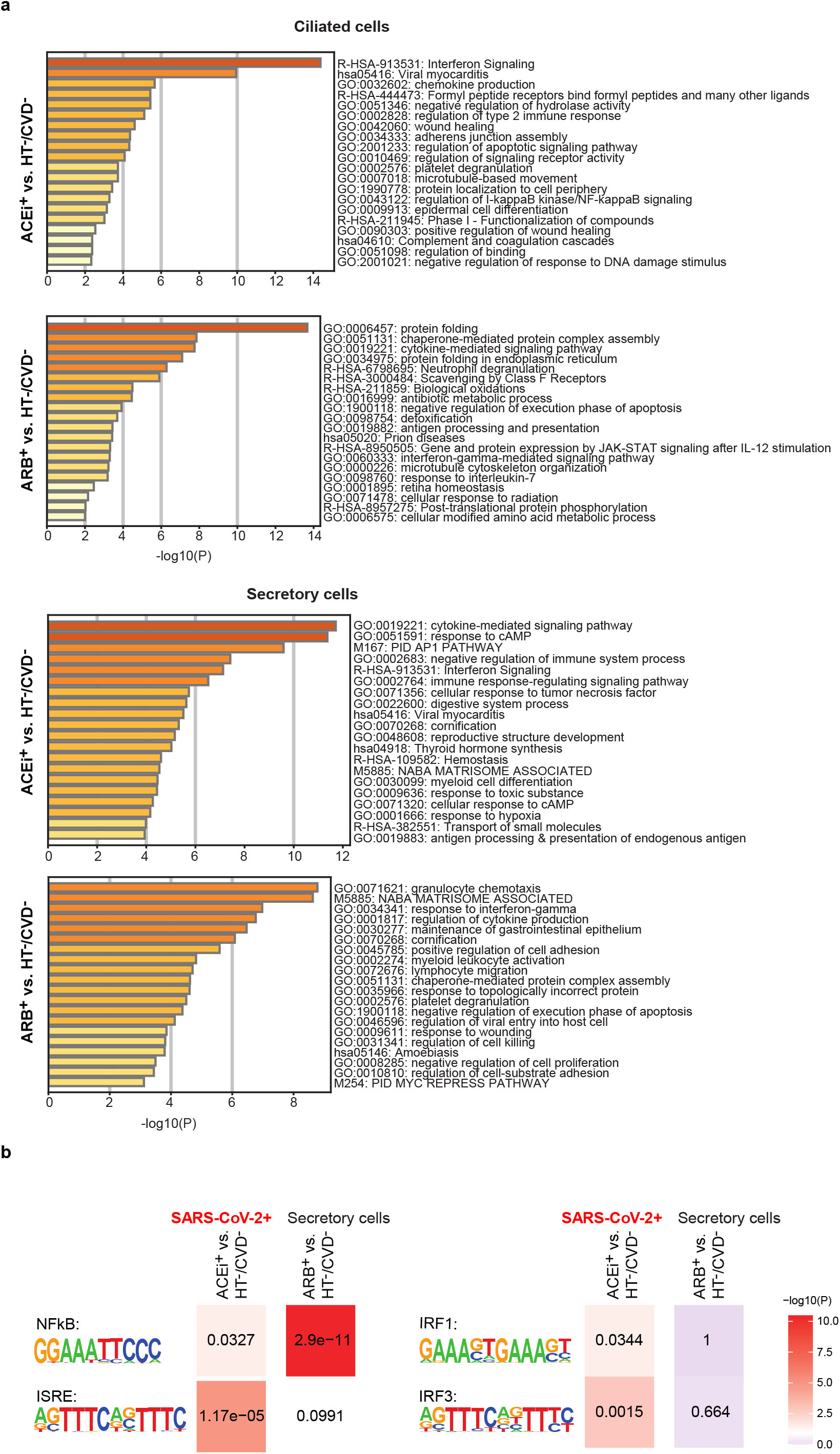
Pathway and transcription factor binding motif enrichment analysis. (a) Pathway enrichment analysis of gene sets (compare Figure 3a) specific to the condition indicated to the right of each panel in ciliated (two upper panels) and secretory cells (two lower panels) in COVID-19 patients. (b) Heatmap showing the p-values of a motif-enrichment analysis for the gene sets upregulated in secretory cells of ACEi+ or ARB+ vs. HT-/CVD-COVID-19 patients. Linear p-values are indicated as labels, log10 p-values are used for color-coding. The patient numbers for deriving the different sets were: SARS-CoV-2+ HT-/CVD-: 8; SARS-CoV-2+ ACEi+: 10; SARS-CoV-2+ ARB+: 15.

**Extended Data Figure 6:**
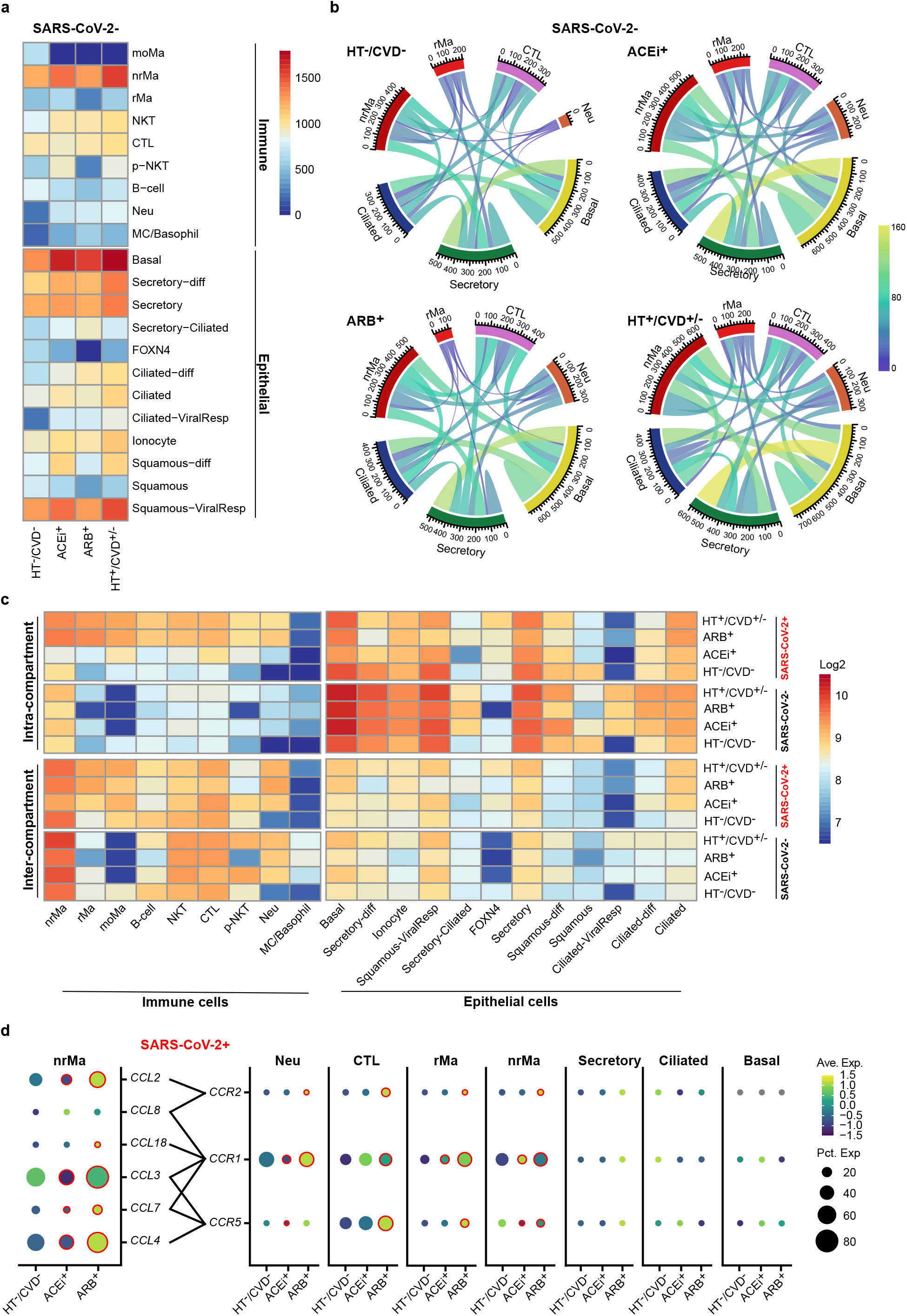
Cell-cell interactions in SARS-CoV-2 negative patients. (a) Heatmap depicting the total number of interactions per cell type across the different SARS-CoV-2-patient conditions. Scaled by the number of identified interactions. (b) Circos plots of highly interactive cells (basal, secretory, ciliated, CTL, Neu, nrMa, and rMa). Scaled by the number of identified interactions. (c) Heatmap showing interactions per cell type categorized as inter-/intra-compartment interactions for SARS-CoV-2 positive and negative cohorts. Log2 scaling of the number of identified interactions. (d) Dotplot showing the expression profile of the different immune modulatory factors by the highly interactive cells. Expression levels are color coded; the percentage of cells expressing the respective gene is size coded. Significantly altered expression (Benjamini–Hochberg adjusted two-tailed, negative-binomial *P* < 0.05) in ACEi+ versus HT-/CVD- (circles around ACEi+) and ARB+ versus HT-/CVD- (circles around ARB+) is marked by a red circle. Ave. Exp. = average expression, Pct. Exp. = percentage of cells expressing the gene. The patient numbers for deriving the different sets were: SARS-CoV-2-HT-/CVD-: 6; SARS-CoV-2-ACEi+: 6; SARS-CoV-2-ARB+: 4; SARS-CoV-2-HT+/CVD±: 10; SARS-CoV-2+ HT-/CVD-: 7; SARS-CoV-2+ ACEi+: 10; SARS-CoV-2+ ARB+: 15; SARS-CoV-2+ HT+/CVD±: 25.

**Extended Data Figure 7:**
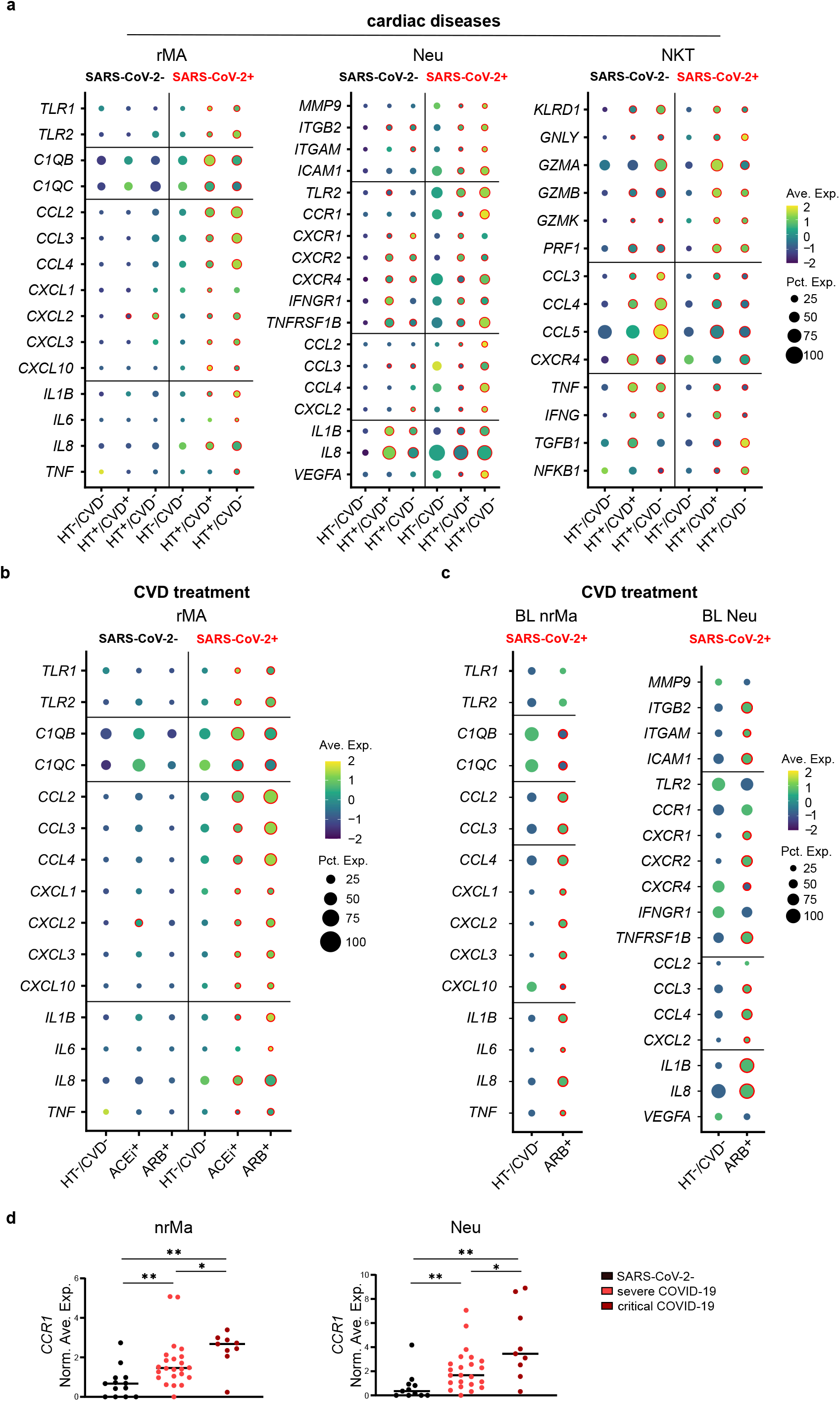
Altered immune response in COVID-19 by ACEi/ARB treatment in relation to disease severity. (a) Dotplot depicts significant gene expression changes of pro-inflammatory mediators, and receptors in rMa, Neu, and NKT of hypertensive patients with (HT+/CVD+, n (SARS-CoV-2-/+) = 4/4) or without an additional CVD (HT+/CVD-, n (SARS-CoV-2-/+) = 6/6) compared to HT-/CVD-patients (n (SARS-CoV-2-/+) = 6/6). Red circles indicate Benjamini-Hochberg adjusted two-tailed negative binominal *p*-value<0.05. Samples with no contributing cells per cell type were excluded from analysis. (b) Alterations of gene expression in rMa of hypertensive patients treated either with ARB+ (n (SARS-CoV-2-/+) = 4/15), or ACEi+ (n (SARS-CoV-2-/+) = 6/10) in comparison to HT-/CVD-patients (n (SARS-CoV-2-/+) = 6/6). (c) Significant changes of gene expression in nrMa and Neu obtained from the bronchial lavage of the COVID-19 HT+/CVD+/ARB+ patient (BIH-SCV2-30) and the HT-/CVD-patient (BIH-SCV2-25). (d) Average gene expression of *CCR1* in nrMa and Neu of SARS-CoV-2 negative (n=11 Neu, n=13 nrMa), severe COVID-19 (n=24 Neu/nrMa), and critical COVID-19 (n=9 Neu/nrMa) patients. Mann-Whitney U-test: ∗*p*-value<0.05, ∗∗*p*-value<0.005, ∗∗∗*p*-value<0.0005. Ave. Exp. = average expression, Pct. Exp. = percentage of cells expressing the gene. BL = bronchial lavage.

## References

1. Grasselli, G. et al. Risk Factors Associated With Mortality Among Patients With COVID-19 in Intensive Care Units in Lombardy, Italy. JAMA Intern Med, E1–E11 (2020).

2. Gupta, S. et al. Factors Associated With Death in Critically Ill Patients With Coronavirus Disease 2019 in the US. JAMA Intern Med, E1-E12 (2020).

3. Danaei, G. et al. National, regional, and global trends in systolic blood pressure since 1980: systematic analysis of health examination surveys and epidemiological studies with 786 country-years and 5.4 million participants. Lancet 377, 568–577 (2011).

4. Huang, S. et al. COVID-19 patients with hypertension have more severe disease: a multicenter retrospective observational study. Hypertens Res 43, 824–831 (2020).

5. Mancia, G., Rea, F., Ludergnani, M., Apolone, G. & Corrao, G. Renin-Angiotensin-Aldosterone System Blockers and the Risk of Covid-19. N Engl J Med 382, 2431–2440 (2020).

6. Gao, C. et al. Association of hypertension and antihypertensive treatment with COVID-19 mortality: a retrospective observational study. Eur Heart J 41, 2058–2066 (2020).

7. Hoffmann, M. et al. SARS-CoV-2 Cell Entry Depends on ACE2 and TMPRSS2 and Is Blocked by a Clinically Proven Protease Inhibitor. Cell 181, 271–280 (2020).

8. Paz Ocaranza, M. et al. Counter-regulatory renin-angiotensin system in cardiovascular disease. Nat Rev Cardiol 17, 116–129 (2020).

9. Romero, C.A., Orias, M. & Weir, M.R. Novel RAAS agonists and antagonists: clinical applications and controversies. Nat Rev Endocrinol 11, 242–252 (2015).

10. Soler, M.J., Barrios, C., Oliva, R. & Batlle, D. Pharmacologic modulation of ACE2 expression. Curr Hypertens Rep 10, 410–414 (2008).

11. Vaduganathan, M. et al. Renin-Angiotensin-Aldosterone System Inhibitors in Patients with Covid-19. N Engl J Med 382, 1653–1659 (2020).

12. Jarcho, J.A., Ingelfinger, J.R., Hamel, M.B., D’Agostino, R.B. Sr. & Harrington, D.P. Inhibitors of the Renin-Angiotensin-Aldosterone System and Covid-19. N Engl J Med 382, 2462–2464 (2020).

13. Reynolds, H.R. et al. Renin-Angiotensin-Aldosterone System Inhibitors and Risk of Covid-19. N Engl J Med 382, 2441–2448 (2020).

14. Dinh, Q.N., Drummond, G.R., Sobey, C.G. & Chrissobolis, S. Roles of inflammation, oxidative stress, and vascular dysfunction in hypertension. Biomed Res Int 2014, 406960 (2014).

15. Jayedi, A. et al. Inflammation markers and risk of developing hypertension: a meta-analysis of cohort studies. Heart 105, 686–692 (2019).

16. Chua, R.L. et al. COVID-19 severity correlates with airway epithelium-immune cell interactions identified by single-cell analysis. Nat Biotechnol, 970-979 (2020).

17. Liao, M. et al. Single-cell landscape of bronchoalveolar immune cells in patients with COVID-19. Nat Med 26, 842–844 (2020).

18. Richardson, S. et al. Presenting Characteristics, Comorbidities, and Outcomes Among 5700 Patients Hospitalized With COVID-19 in the New York City Area. JAMA, 2052-2059 (2020).

19. Liu, S. et al. Clinical characteristics and risk factors of patients with severe COVID-19 in Jiangsu province, China: a retrospective multicentre cohort study. BMC Infect Dis 20, 1–9 (2020).

20. Benelli, G. et al. SARS-COV-2 comorbidity network and outcome in hospitalized patients in Crema, Italy. preprint at https://doi.org/10.1101/2020.04.14.20053090 (2020).

21. Kurth, F. et al. Studying the pathophysiology of coronavirus disease 2019: a protocol for the Berlin prospective COVID-19 patient cohort (Pa-COVID-19). Infection 48, 619–626 (2020).

22. Lukassen, S. et al. SARS-CoV-2 receptor ACE2 and TMPRSS2 are primarily expressed in bronchial transient secretory cells. EMBO J 39, e105114 (2020).

23. Hou, Y.J. et al. SARS-CoV-2 Reverse Genetics Reveals a Variable Infection Gradient in the Respiratory Tract. Cell, 429–446 (2020).

24. Nawijn, M.C. & Timens, W. Can ACE2 expression explain SARS-CoV-2 infection of the respiratory epithelia in COVID-19? Mol Syst Biol 16, e9841 (2020).

25. Park, A. & Iwasaki, A. Type I and Type III Interferons-Induction, Signaling, Evasion, and Application to Combat COVID-19. Cell Host Microbe 27, 870–878 (2020).

26. Taniguchi, K. & Karin, M. NF-kappaB, inflammation, immunity and cancer: coming of age. Nat Rev Immunol 18, 309–324 (2018).

27. Liu, T., Zhang, L., Joo, D. & Sun, S.C. NF-kappaB signaling in inflammation. Signal Transduct Target Ther 2, e17023 17021–17029 (2017).

28. Neufeldt, C.J. et al. SARS-CoV-2 infection induces a pro-inflammatory cytokine response through cGAS-STING and NF-κB. preprint at https://doi.org/10.1101/2020.07.21.212639 (2020).

29. Efremova, M., Vento-Tormo, M., Teichmann, S.A. & Vento-Tormo, R. CellPhoneDB: inferring cell-cell communication from combined expression of multi-subunit ligand-receptor complexes. Nat Protoc 15, 1484–1506 (2020).

30. Guan, W.J. et al. Comorbidity and its impact on 1590 patients with COVID-19 in China: a nationwide analysis. Eur Respir J 55, 1–14 (2020).

31. Sanyaolu, A. et al. Comorbidity and its Impact on Patients with COVID-19. SN Compr Clin Med, 1–8 (2020).

32. Zhang, P. et al. Association of Inpatient Use of Angiotensin-Converting Enzyme Inhibitors and Angiotensin II Receptor Blockers With Mortality Among Patients With Hypertension Hospitalized With COVID-19. Circ Res 126, 1671–1681 (2020).

33. Chung, M.K. et al. SARS-CoV-2 and ACE2: The biology and clinical data settling the ARB and ACEI controversy. EBioMedicine 58, 102907 (2020).

34. Mehta, N. et al. Association of Use of Angiotensin-Converting Enzyme Inhibitors and Angiotensin II Receptor Blockers With Testing Positive for Coronavirus Disease 2019 (COVID-19). JAMA Cardiol, 1–8 (2020).

35. Nyambuya, T.M., Dludla, P.V., Mxinwa, V. & Nkambule, B.B. T-cell activation and cardiovascular risk in adults with type 2 diabetes mellitus: A systematic review and meta-analysis. Clin Immunol 210, 108313 108311–108312 (2020).

36. Kintscher, U. et al. Plasma Angiotensin Peptide Profiling and ACE2-Activity in COVID-19 Patients treated with Pharmacological Blockers of the Renin Angiotensin System. Hypertension, 1–4 (2020).

## Online Methods References

37. Williams, B. et al. 2018 ESC/ESH Guidelines for the management of arterial hypertension: The Task Force for the management of arterial hypertension of the European Society of Cardiology and the European Society of Hypertension: The Task Force for the management of arterial hypertension of the European Society of Cardiology and the European Society of Hypertension. J Hypertens 36, 1953–2041 (2018).

38. Aylward, B., Liang, W. & WHO-China-Joint-Mission Report of the WHO-China Joint Mission on Coronavirus Disease 2019 (COVID-19). published on the WHO website, document can be found at https://www.who.int/publications-detail/report-of-the-who-china-joint-mission-on-coronavirus-disease-2019-(covid-19) (page 12) (2020).

39. Young, M. & Behjati, S. SoupX removes ambient RNA contamination from droplet based single-cell RNA sequencing data. preprint at https://doi.org/10.1101/303727 (2020).

40. Plasschaert, L.W. et al. A single-cell atlas of the airway epithelium reveals the CFTR-rich pulmonary ionocyte. Nature 560, 377–381 (2018).

41. Vieira Braga, F.A. et al. A cellular census of human lungs identifies novel cell states in health and in asthma. Nat Med 25, 1153–1163 (2019).

42. Travaglini, K.J. et al. A molecular cell atlas of the human lung from single cell RNA sequencing. preprint at https://doi.org/10.1101/742320 (2019).

43. Yu, G., Wang, L.G., Han, Y. & He, Q.Y. clusterProfiler: an R package for comparing biological themes among gene clusters. OMICS 16, 284–287 (2012).

44. Gu, Z., Gu, L., Eils, R., Schlesner, M. & Brors, B. circlize Implements and enhances circular visualization in R. Bioinformatics 30, 2811–2812 (2014).

45. Moerman, T. et al. GRNBoost2 and Arboreto: efficient and scalable inference of gene regulatory networks. Bioinformatics 35, 2159–2161 (2019).

46. Aibar, S. et al. SCENIC: single-cell regulatory network inference and clustering. Nat Methods 14, 1083–1086 (2017).

47. Corman, V.M. et al. Detection of 2019 novel coronavirus (2019-nCoV) by real-time RT-PCR. Euro Surveill 25, 1–8 (2020).

48. Binder, M. et al. Molecular mechanism of signal perception and integration by the innate immune sensor retinoic acid-inducible gene-I (RIG-I). J Biol Chem 286, 27278–27287 (2011).

49. Pfaender, S. et al. LY6E impairs coronavirus fusion and confers immune control of viral disease. Nat Microbiol (2020).

50. Heinz, S. et al. Simple combinations of lineage-determining transcription factors prime cis-regulatory elements required for macrophage and B cell identities. Mol Cell 38, 576–589 (2010).

